# Genome-wide fine-mapping improves identification of causal variants

**DOI:** 10.1101/2024.07.18.24310667

**Authors:** Yang Wu, Zhili Zheng, Loic Thibaut, Tian Lin, Qian Feng, Hao Cheng, Loic Yengo, Michael E. Goddard, Naomi R. Wray, Peter M. Visscher, Jian Zeng

## Abstract

Fine-mapping refines genotype-phenotype association signals to identify causal variants underlying complex traits. However, current methods typically focus on individual genomic loci and do not account for the global genetic architecture. Here, we demonstrate the advantages of performing genome-wide fine-mapping (GWFM) with functional annotations and develop methods to facilitate GWFM. In simulations and real data analyses, GWFM outperforms current methods across multiple metrics, including error control, mapping power, resolution, precision, replication rate, and trans-ancestry phenotype prediction. Across 48 complex traits, we identify credible sets that collectively explain 18% of the SNP-based heritability 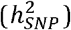 on average, with 30% credible sets located outside genome-wide significant loci. Leveraging the genetic architecture estimated from GWFM, we predict that fine-mapping over 50% of 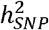 would require an average of 2 million samples. Finally, as proof-of-principle, we highlight a known causal variant at FTO for body mass index and identify novel missense causal variants for schizophrenia and Crohn’s disease.

## Introduction

Despite the success of genome-wide association studies (GWAS) in identifying trait-associated variants^1^, the causal variants underlying complex traits remain unresolved due to extensive linkage disequilibrium (LD) between SNPs^2^ and the polygenic nature of these traits^3,4^. Statistical fine-mapping, often employing a Bayesian mixture model (BMM) that jointly fit multiple SNPs, offers a direct approach to identifying candidate causal variants^5^. However, current fine-mapping methods focus on genome-wide significant loci only (e.g., 1-2 Mb windows centred on lead SNPs after LD clumping^6-12^) or consider one genomic region at a time (e.g., a LD block^13^), in isolation from the rest of the genome.

While widely used, region-specific analysis has several limitations. First, restricting fine-mapping to GWAS loci alone, which often explain only a small fraction of the total genetic variance^14,15^, omits meaningful signals that have not yet reached the stringent GWAS significance threshold. Second, the prior probability of association can be influential but is often conservatively predetermined (e.g., as the inverse of the number of SNPs in the region^6,7,16^) due to challenges in estimating the genetic architecture within a region. Third, fine-mapping can benefit from incorporating functional genomic annotations^10-12,17^, but region-specific methods often estimate functional priors separately before performing functionally-informed fine-mapping^10-12,17^, rather than jointly modelling GWAS data and functional annotations. Finally, none of the current methods estimate the power of identifying the causal variants for a trait, which is critical to inform the experimental design of prospective studies (such a power analysis is available in GWAS^18^ but absent in fine-mapping).

These limitations can be addressed with genome-wide fine-mapping (GWFM) analysis. Genome-wide Bayesian mixture models (GBMMs), extensively used for predicting breeding values in agriculture^19-21^ and complex trait phenotypes in humans^22-24^, have recently emerged as a method for GWFM^25,26^. Unlike conventional GWAS and region-specific fine-mapping approaches, GBMMs jointly fit genome-wide SNPs in the model, simultaneously estimating the genetic architecture and functional priors through “information borrowing”^24,25^. For example, SNPs sharing the same functional annotation across the genome collectively prioritize that annotation based on their aggregated evidence of trait association, which, in turn, informs the estimation of individual SNP effects. This learning process is often performed iteratively using Markov chain Monte Carlo (MCMC) sampling, leading to posterior inference with superior asymptotic accuracy compared to variational inference^27,28^, though MCMC sampling can be computationally intensive with high-density SNPs. Moreover, GBMMs estimate polygenicity and the distribution of variant effect sizes^3,20,22-24,29^, enabling the prediction of power for prospective studies with larger sample sizes. However, relevant theory and methods have not yet been developed.

In this study, we comprehensively evaluated the performance of GWFM analysis using SBayesRC^24^, a state-of-the-art GBMM that enables efficient MCMC-based fitting of all common SNPs with functional annotations. Through extensive simulations under various genetic architectures, we compared methods using multiple metrics, including posterior inclusion probability (PIP) calibration, fine-mapping power, mapping precision, credible set size, replication rate in independent samples, and out-of-sample prediction accuracy using fine-mapped variants. We developed an LD-based method to construct local credible sets (*α*-LCSs) for GBMM, each capturing a causal variant with posterior probability *α*, and estimated the proportion of SNP-based heritability 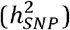 explained by these LCSs. To quantify overall fine-mapping uncertainty, we provided a global credible set (*α*-GCS) that captures *α*% of all causal variants for the trait. Leveraging the genetic architecture estimated from SBayesRC, we further developed a method to predict fine-mapping power and the 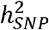 explained by identified variants in prospective studies, enabling estimation of the minimal sample size required to capture a desired proportion of causal variants or 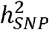. Finally, we applied SBayesRC to 599 complex traits and diseases with 13 million SNPs and compared the fine-mapping results for 48 well-powered traits, primarily using data from the UK Biobank^30^ (UKB).

## Results

### Method overview

We selected SBayesRC as the method for GWFM (**Fig. 1**) because it outperforms other GBMMs in polygenic prediction^24^. SBayesRC is a multi-component mixture model that simultaneously analyzes all SNPs across approximately independent LD blocks^13,31^, using a hierarchical prior to borrow information from functional annotations across the genome for fine-mapping within blocks, where both annotation and SNP effects are jointly estimated from the data (**Methods**). Unlike existing fine-mapping methods that focus on specific GWAS loci, SBayesRC accounts for long-range LD (variable LD block sizes with a minimum of 1 Mb, including the major histocompatibility complex (MHC) region) and maps causal signals over the entire genome (**Supplementary Table 1**). To optimize fine-mapping performance, we heuristically estimated the number of mixture components, adapted a tempered Gibbs sampling algorithm^32^ to improve mixing properties, and implemented a method^33^ to assess MCMC convergence using multiple independent chains (**Methods**). Further methodological differences between our approach and existing methods are discussed in **Supplementary Note 1**.

**Figure 1.**
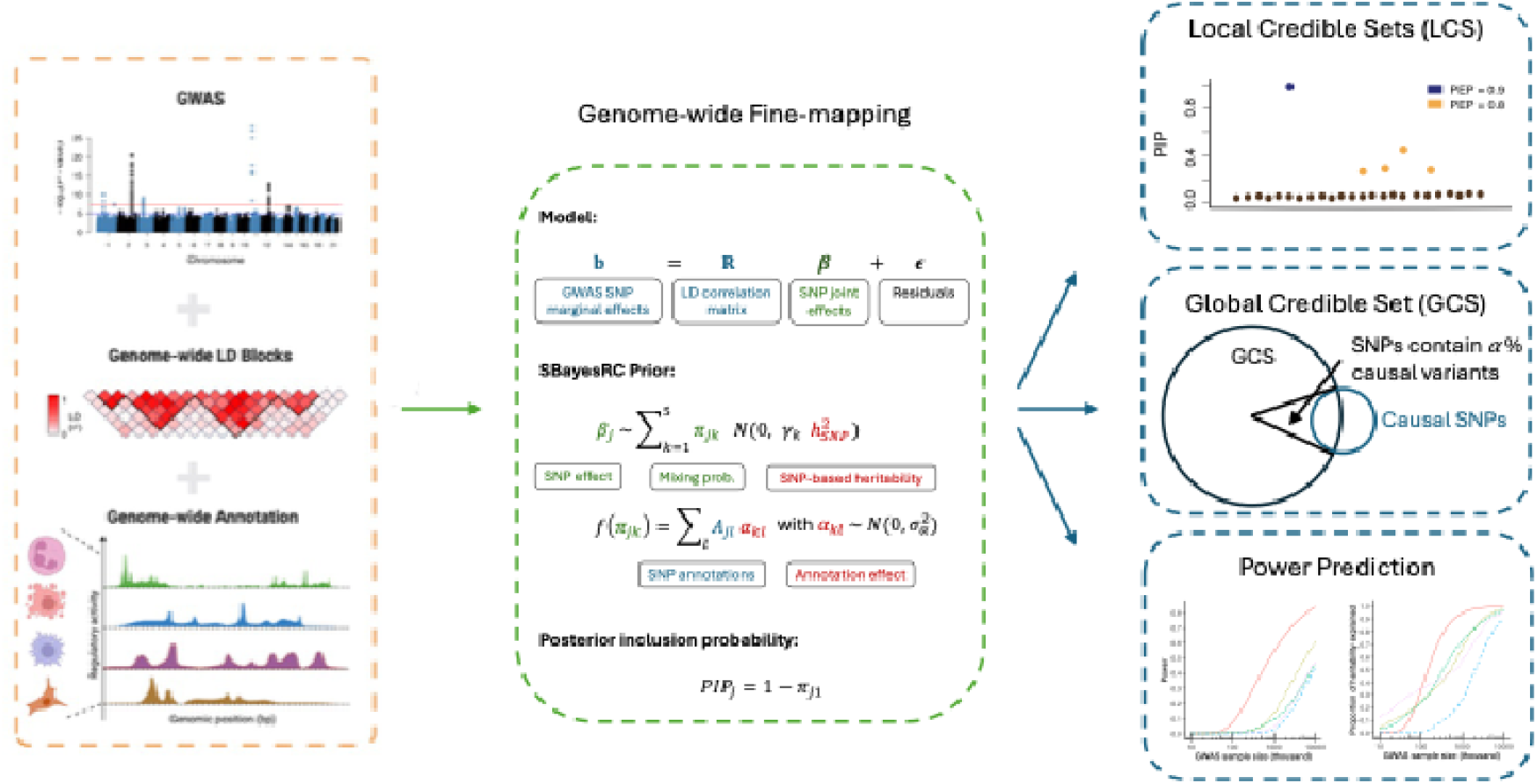
Schematic overview of genome-wide fine-mapping (GWFM) analysis using GBMM. GBMM utilizes GWAS summary statistics and genome-wide LD reference to fine-map candidate causal variants for complex traits, incorporating functional annotations when available. Unlike region-specific fine-mapping approaches, GBMM maps causal signals across the entire genome and provides greater flexibility in modelling the distribution of causal effects by estimating priors using genome-wide SNPs (**Table S1**). Different GBMMs share the same fitted model but differ in prior specification for SNP effects. As shown in the middle box, SBayesRC assumes a mixture prior consisting of a point mass at zero and multiple normal distributions with variance 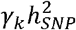, where *γ*_*k*_ = [0,1^−5^,1^−4^,1^−3^,1^−2^]’. The mixing probability for SNP *j* in distribution *k* (*π*_*jk*_) is modelled as a linear combination of SNP annotations through a probit link. The color scheme is as follows: blue denotes input data, green denotes SNP-specific parameters, and red denotes global parameters shared across SNPs. The GBMM output includes SNP PIPs, local and global credible sets, and prediction of fine-mapping power for future studies. The illustration in the left box was created with BioRender.com.

To capture the uncertainty in identifying causal variants driven by LD, we developed an LD-based method for constructing *α*-LCSs (**Methods; Supplementary Fig. 1**). Our approach forms an *α* -LCS for each candidate causal variant with a leading PIP by grouping it with SNPs in LD (r^2^ > 0.5) until their combined PIPs sum to *α*. To refine the LCSs, we further filter them based on the posterior 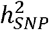 enrichment probability (PEP > 0.7) to ensure that the selected LCS explains more 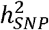 than a random set of SNPs with the same size, using MCMC samples of SNP effects (Methods). Additional justification for the r^2^ and PEP thresholds is provided in **Supplementary Note 2** and **Supplementary Fig. 2-3**.

Since SBayesRC estimates the genetic architecture of the trait, this framework lends itself to quantifying overall uncertainty in fine-mapping and predicting power for future GWFM studies. The overall uncertainty in the current study is captured by our *α*-GCS, which is expected to cover *α* % of all causal variants for the trait, with the size of *α*-GCS reflecting power and genetic architecture (**Methods**). Fine-mapping power prediction for prospective studies is achieved by deriving the sampling distribution of PIP given a sample size (**Methods**). This method allows us to estimate the minimal sample size required to achieve the desired power to identify all causal variants or those explaining a specific proportion of 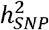. Although we focused on European ancestry in this study, our power prediction method is, in principle, applicable to other ancestries; however, the genetic architecture would need to be estimated using ancestry-specific GWAS data, as it may vary across populations. Our fine-mapping power prediction method is analytically tractable and available as a publicly accessible online tool (https://sbayes.pctgplots.cloud.edu.au/shiny/power/).

### Calibration of fine-mapping methods under various genetic architectures

We performed extensive genome-wide simulations to evaluate SBayesRC for GWFM, comparing to FINEMAP^7^, SuSiE^6^, FINEMAP-inf^8^, SuSiE-inf^34^, PolyFun+SuSiE^10^, and another two GWFM methods, SBayesR^22^ and SBayesC (two-component SBayesR). For a fair comparison, we defined fine-mapping regions consistently across all methods using independent LD blocks and applied the default parameter settings recommended for each method (**Methods**). Using 100,000 individuals with ∼1 million HapMap3 SNPs from the UKB^30^, we simulated three genetic architectures: 1) a sparse architecture with 1% randomly selected causal SNPs explaining 50% of phenotypic variance, 2) a large-effects architecture where 10 random causal variants contributed 10% of phenotypic variance and the rest 40%, and 3) an LD-and-MAF-stratified (LDMS) architecture with causal variants sampled from high LD score and high minor allele frequency (MAF) SNPs (**Methods**).

Results showed that GWFM methods generally performed better in terms of PIP calibration compared to region-specific methods, with PIPs from SBayesRC closely aligning with true probabilities of causality (measured by true discovery rate, TDR) across all genetic architectures (**Fig. 2a,d,g**). In contrast, SuSiE and FINEMAP exhibited notable inflation in high-PIP SNPs (**Fig. 2b,e,h**), indicating poor control of false discovery rate (FDR = 1-TDR). While SuSiE-inf and FINEMAP-inf performed reasonably well under the sparse and large-effects architectures (despite some deflation in low-PIP SNPs), their PIPs failed to track TDR accurately under the LDMS architecture, where causal variants were not randomly distributed (**Fig. c,f,i**). Polyfun+SuSiE, when incorporating the same LD/MAF annotations as SBayesRC, showed improved performance over SuSiE and FINEMAP but still struggled with FDR control (**Fig. 2i**). The alternative GWFM methods, SBayesC and SBayesR, were inferior to SBayesRC under the large-effects and LDMS architectures, respectively, highlighting the importance of using multiple mixture components and integrating informative genomic annotations for robust calibration.

**Figure 2.**
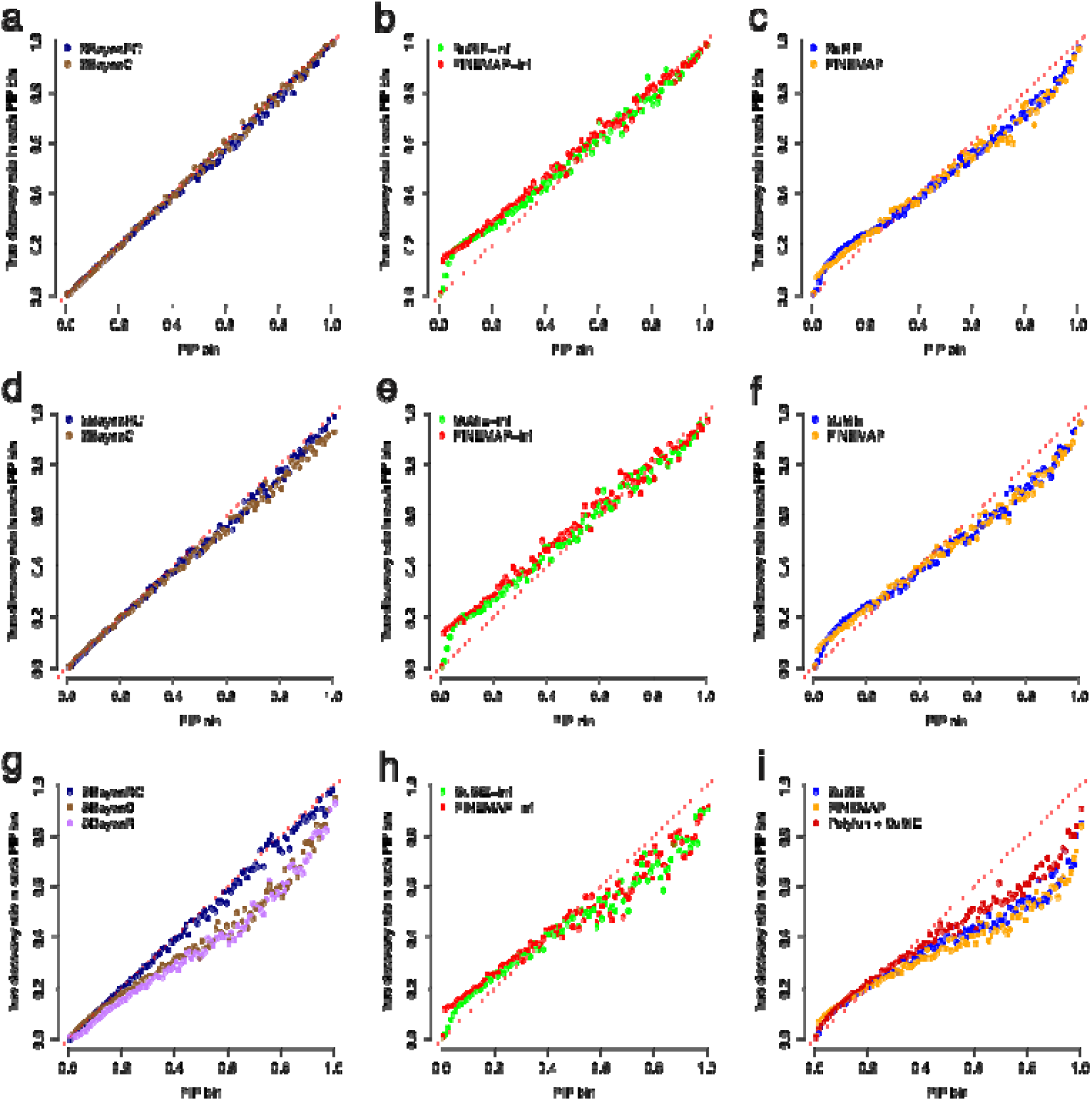
Comparison in the calibration of PIP between GWFM and existing fine-mapping methods under simulations with various genetic architectures. Shown are relationships between PIP and the true discovery rate (TDR) across 100 PIP bins. Left column: SBayesC, SBayesR, and SBayesRC; middle column: SuSiE-inf and FINEMAP-inf; right column: SuSiE, FINEMAMP, and Polyfun + SuSiE. Top row: sparse genetic architecture; middle row: large-effects genetic architecture; Bottom row: LDMS architecture.

### Assessing mapping power, resolution, and precision via simulations

We first compared the power of different fine-mapping strategies, GWFM and GWAS loci-based fine-mapping, by evaluating the proportion of causal variants captured by *α*-LCSs. For a fair comparison, GWAS loci-based fine-mapping used the same PIP estimates as GWFM with SBayesRC but was restricted to 2Mb regions around GWAS lead SNPs (P-value < 5×10^−8^). As expected, GWFM was significantly more powerful, with 46%-61% improvement across genetic architectures (**Fig. 3a-c**). This highlights the advantage of considering the entire genome, including regions not genome-wide significant in GWAS, in fine-mapping.

**Figure 3.**
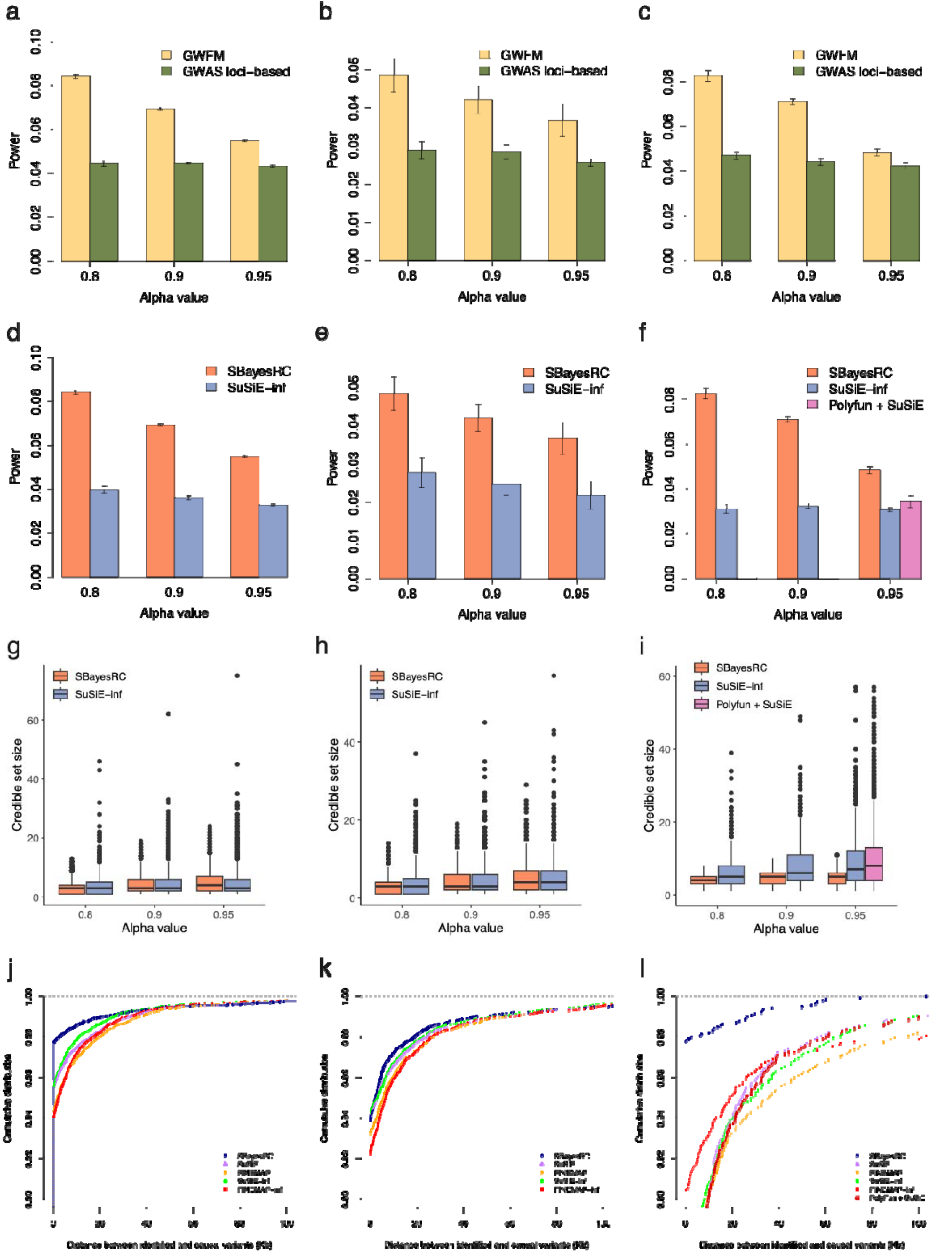
Comparison in mapping power, resolution, and precision between methods based on simulations. Panels (a-c) compare mapping power between GWFM and GWAS loci-based strategies at the same confidence level (*α*) for LCSs. For a fair comparison, both strategies used the same PIP estimates from SBayesRC, whereas GWAS loci-based fine-mapping constructed α-LCSs restricted to 2Mb regions around GWAS lead SNPs (P-value < 5×10^−8^). Panels (d-f) compare mapping power across different methods, including SBayesRC, SuSiE-inf, Polyfun + SuSiE. Power was quantified as the proportion of simulated causal variants included in the credible sets reported by each method. Panels (g-i) compare mapping resolution across SBayesRC, SuSiE-inf, and Polyfun + SuSiE, measured by the credible set size. Panels (j-l) compare mapping precision across different methods, measured by the physical distance between the causal variants and the SNPs identified at PIP ≥ 0.9 in each method. Columns correspond to simulations under the sparse genetic architecture (left column), large-effects genetic architecture (middle column), and LDMS genetic architecture (right column).

Next, we compared different fine-mapping methods, focusing on SBayesRC and SuSiE-inf, as SuSiE-inf had the best PIP calibration and least FDR inflation among the region-specific methods. Our results showed that SBayesRC was significantly more powerful (**Fig. 3d-f**), with similar TDR (**Supplementary Fig. 4a-c**) but smaller LCS sizes at the same *α* threshold, indicating superior mapping resolution (**Fig. 3g-i**). For *α* =0.9, SBayesRC outperformed SuSiE-inf by up to 194% increase in power and 21% reduction in average LCS size across the three genetic architectures. Polyfun+SuSiE, which incorporated LD/MAF annotations through a stepwise analysis, performed slightly better than SuSiE-inf under the LDMS architecture but remained inferior to SBayesRC. Mapping precision was assessed as the distance from an identified variant (PIP > 0.9) to the nearest causal variant. Under the sparse architecture, 98% of SNPs identified by SBayesRC were causal, and 99% of SNPs with PIP > 0.9 were within 18kb of a causal variant, leading to a 2% increase in TDR and a 68% reduction in distance compared to other methods (**Fig. 3j**). The advantage of SBayesRC was even more pronounced under the large-effects and LDMS architecture (**Fig. 3k-l**), likely due to its ability to differentially weigh SNPs based on LD and MAF annotations, thereby improving causal variants identification.

Furthermore, we used these simulations to evaluate GCS (**Supplementary Fig. 5**) and conducted sensitivity analyses to confirm the robustness of SBayesRC to missing annotations (**Supplementary Fig**. 6), unobserved causal variants (**Supplementary Fig. 7**), and the chosen value of LD matrix factorization parameter (**Supplementary Fig. 8**). To understand why GWFM had higher power, we investigated power gains in causal variants with different LD and MAF properties (**Supplementary Fig. 9**) and assessed SBayesRC within each LD block separately (**Supplementary Fig. 10**). These results are discussed in **Supplementary Note 3**. Overall, these simulation results suggested that SBayesRC is a reliable method for GWFM and can substantially improve fine-mapping performance.

### Assessing replication rate, effect size estimation, and prediction accuracy in real data

In real data analysis, direct evaluation of mapping power and precision is challenging since true causal variants are often unknown. Thus, we assessed methods using metrics that do not require knowledge of causal variants. First, we evaluated replication rate by calculating the proportion of variants with PIP > 0.9 in a GWAS sample that were also identified in an independent replication sample at the same or a lower PIP threshold. Assuming no systematic confounding, a higher replication rate suggests that the method identifies more genuine causal variants, as false positives are less likely to replicate. Using UKB height data (n=100,000) as a GWAS sample, SBayesRC achieved the highest replication rate in each PIP threshold, improving by 1% (compared to SuSiE-inf) to 11% (compared to FINEMAP) at PIP > 0.9 when replication *n*=100,000 (**Fig. 4a**). When replication sample size was doubled, the improvement increased to 4% compared to SuSiE-inf and 19% compared to FINEMAP (**Supplementary Fig. 11**).

**Figure 4.**
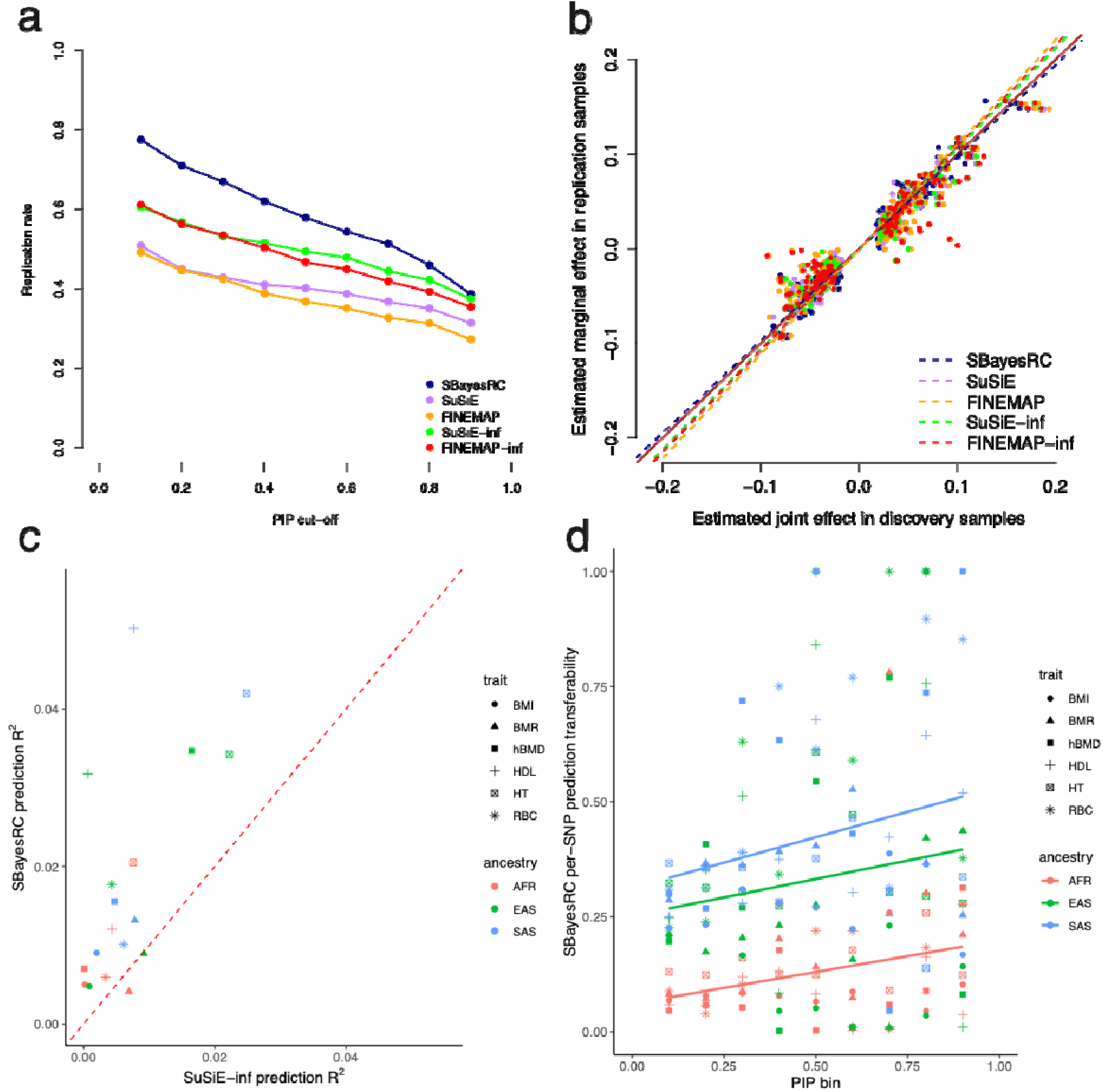
Comparison of independent sample replication, effect size estimation bias, and prediction accuracy using fine-mapped variants across fine-mapping methods in UK biobank traits. (a) Replication rate of discovery using different methods at a given PIP threshold in the replication sample (x-axis) for height in the UKB. (b) Regression of the estimated marginal effect sizes in replication samples on the estimated joint effect sizes in discovery samples using different fine-mapping methods. Dash lines show the regression slopes, where values closer to one indicate less bias, as the marginal effect estimated in the independent replication sample is an unbiased estimate to the true effect. The brown solid line is y=x. (c) Comparison of trans-ancestry prediction accuracy using fine-mapped variants (PIP > 0.9) from SBayesRC and SuSiE-inf, based on the analysis using samples of European ancestry for GWAS and the other ancestries for validation across 6 UKB traits. BMI: body mass index; BMR: base metabolic rate; hBMD: heel bone mineral density; HDL: high-density lipoprotein level; HT: height; RBC: red blood cell count. AFR: African ancestry; EAS: East Asian ancestry; SAS: South Asian ancestry. (d) Relationship between trans-ancestry prediction transferability and PIP estimates in the European ancestry. The transferability was computed as non-EUR-R^2^/ EUR-R^2^. The solid lines are regression lines across traits in each ancestry.

We assessed bias in effect size estimates of putative causal variants through regressing their marginal effect sizes from the replication sample on the joint effect sizes estimated from the GWAS sample, expecting a regression slope of one for unbiased estimation. In the UKB height analysis, SBayesRC produced the minimal bias, with a regression slope of 0.98, outperforming all the other methods (**Fig. 4b**). This likely reflects the capacity of SBayesRC to estimate genetic architecture genome-wide, whereas other methods estimate genetic architecture locally or use preset parameters.

Given that common causal variants and their effect sizes are mostly shared across ancestries^35,36^, knowing the causal variants should improve trans-ancestry prediction in complex traits. Therefore, we used fine-mapped variants and their posterior effect sizes from UKB European (EUR) ancestry individuals to predict phenotypes in African (AFR), East Asian (EAS), and South Asian (SAS) ancestry groups. We selected 6 complex traits that had at least 50 SNPs with PIP > 0.9. Compared to SuSiE-inf, SBayesRC improved the trans-ancestry prediction accuracy, with nearly a 10-fold increase in mean relative prediction R^2^ across traits and ancestries (**Fig. 4c**). Similar improvements were observed when using 0.9-LCSs instead of individual SNPs (**Supplementary Fig. 12**) or when comparing to Polyfun+SuSiE with the same functional annotations (**Supplementary Fig. 13**). We further quantified the transferability of fine-mapped SNPs by computing the ratio of per-SNP prediction accuracy in a hold-out EUR sample versus a different ancestry. The result showed that relative prediction accuracy increased with PIP in the EUR GWAS sample (**Fig. 4d**), consistent with a model of shared genetic effects across ancestries.

We validated these advantages of SBayesRC through simulations, confirming its superior power (**Supplementary Fig. 14**), replication rate (**Supplementary Fig. 15a**), effect size estimation (**Supplementary Fig. 15b**), and out-of-sample prediction using fine-mapped variants (**Supplementary Fig. 16**), with further discussions in **Supplementary Note 4**. We also quantified the replication rate in the reverse case where the GWAS sample size was 200,000 but the replication sample size was only 100,000, to mimic the reality that the sample size of replication data is often much smaller than that of discovery, and again observed superior replication rate of our SBayesRC (**Supplementary Fig. 17a**,**b**). These results suggested that SNPs identified by SBayesRC are more likely to be causal, as indicated by higher replication rates, while the improved prediction accuracy likely arises from both better causal variant identification and more accurate effect size estimation.

### Prediction of fine-mapping power and variance explained for future studies

As a unique feature of the GWFM approach, the genetic architecture estimated from SBayesRC provides key information to predict the proportion of causal variants identified (fine-mapping power) and the proportion of 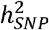 explained by these variants (PHE) in future studies (Methods). To evaluate our approach, we computed the predicted values of power and PHE across varying GWAS sample sizes and projected the outcome of GWFM with SBayesRC onto the prediction for a simulated trait (under the sparse architecture), height^30,37^, high density lipoprotein (HDL)^30,38^, schizophrenia (SCZ)^39,40^, and Crohn’s disease (CD)^30,41^, which represented diverse genetic architectures, using two datasets with different sample sizes for each trait (Fig. 5a-c). Despite some variation across traits, GWFM outcomes were broadly consistent with the theoretical predictions (Fig. 5d,e).

**Figure 5.**
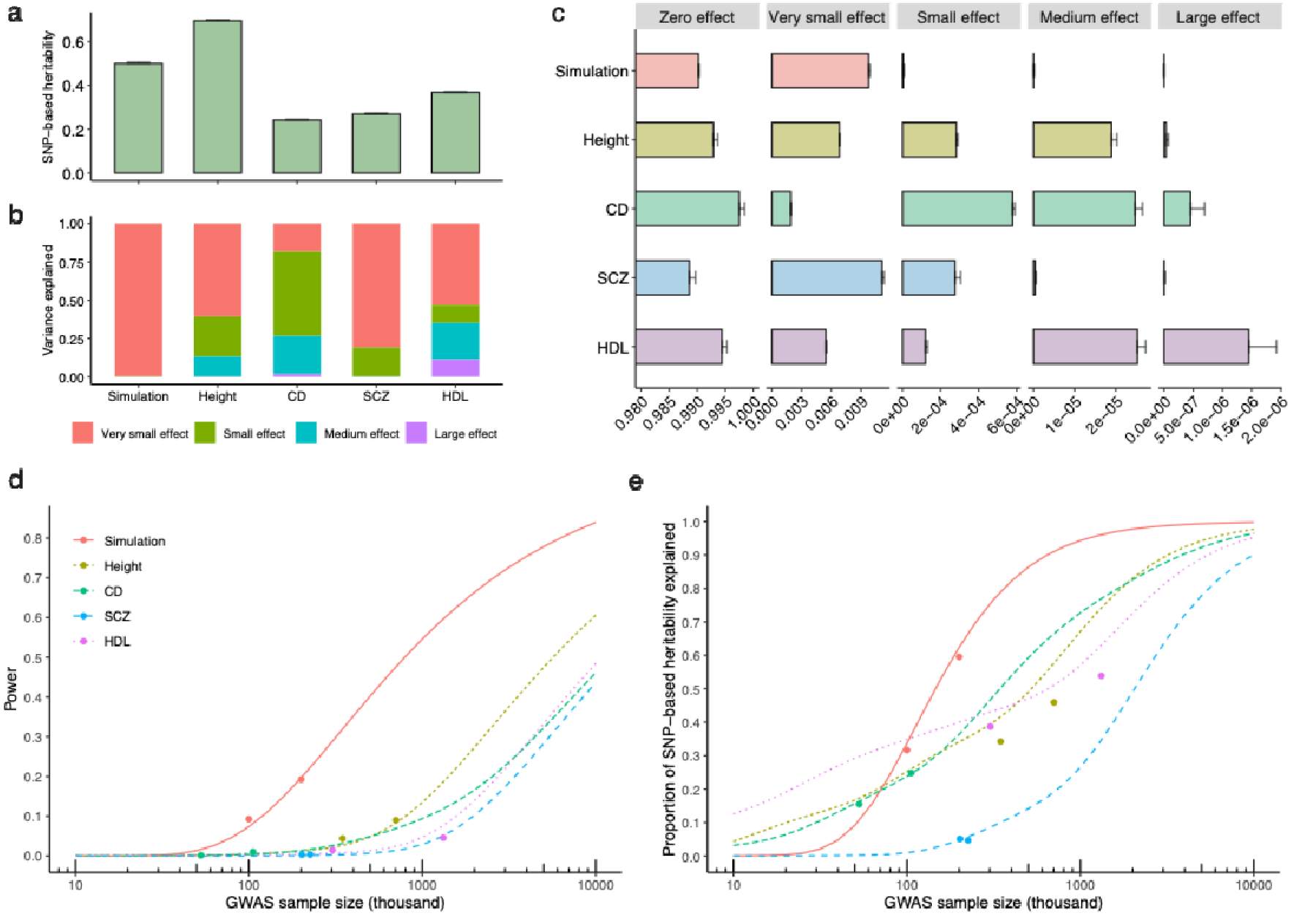
Projection of genome-wide fine-mapping outcomes to the theoretical power prediction in complex traits with diverse genetic architectures. (a) Estimates of SNP-based heritability 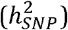 obtained using SBayesRC for height, Crohn’s disease (CD), schizophrenia (SCZ), high density lipoprotein (HDL), and a simulated trait with the sparse genetic architecture. (b) Proportions of 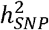 explained by different mixture components in the SBayeRC model. (c) Proportions of SNPs assigned to different mixture components (***π***) based on their estimated effect sizes. The “zero”, “very small”, “small”, “medium” and “large” effect categories correspond to the five variance components in SBayesRC, which explain 0, 0.001, 0.01, 0.1, and 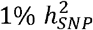, respectively. For the simulated traits, all 10,000 causal variants had small effects sampled from a single normal distribution, leading to all estimated effect sizes being assigned to the very small effect category. (d-e) Theoretical prediction of the power of identifying causal variants (d) and the proportion of 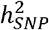 explained by the identified causal variants (e) across different GWAS sample sizes for these traits. Dots show the empirical observations based on 0.9-LCSs identified from SBayesRC. The genetic architecture estimates from (a, c) were used as input data for the fine-mapping power prediction (d-e). To check consistency, two datasets with different GWAS sample sizes were analyzed for each trait, with genetic architecture estimates derived from the dataset with the smaller sample size.

Using the latest GWAS summary statistics from the Psychiatric Genomics Consortium (PGC)^39^, we identified 13 SNPs and 174 LCSs for SCZ, collectively explaining 4.2% of 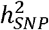 on the liability scale (converted from observed 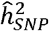 based on ref^42^). These estimates aligned closely with our theoretical prediction, based on the 53,386 cases and 77,258 controls in their study^39^, which is equivalent to a sample size of 228,810 on the liability scale (ref^43^; Methods). For a prospective SCZ study using GWFM, we predict that ∼180k cases (with equal controls and a population prevalence of 0.01) would be required to fine-map 1,000 common causal variants (estimated to be 1.2% of all common causal variants), collectively explaining ∼20% of 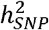(**Fig. 5**). Increasing the sample size to ∼550k cases would allow for the identification of 10% of causal variants explaining 50% of 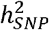, while fine-mapping variants accounting for 80% of 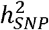would require 1.4 million cases according to our prediction.

For height, based on the UKB data (n=350k), we predicted that with 5 million samples, ∼10,000 variants would be identified with individual PIP > 0.9 or ∼30,000 when considering LCSs, explaining up to 95% of the genetic variance (**Fig. 5**). This prediction is consistent with a recent GWAS of 5 million individuals, which reported 12,111 independently significant SNPs identified from COJO accounting for nearly all of the common 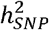 in height^36^.

### Applying genome-wide fine-mapping to a range of complex traits

We applied GWFM with SBayesRC to 599 complex traits (597 UKB traits plus SCZ^39^ and CD^41^) and developed an online resource to query these fine-mapping results (see **Data Availability; Supplementary Table 3**). The 597 UKB traits were selected based on z-score > 4 and high confidence for heritability estimates using LD score regression^44^ (https://zenodo.org/records/7186871). To better capture causal variants, we used 13 million imputed SNPs with functional genomic annotations from Finucane et al.^45^. Here, we focus on 48 well-powered traits, including SCZ, CD^39,41^ and 46 UKB traits measured in the European ancestry individuals (**Methods**).

Across the 48 traits, we identified 1,820 SNPs with PIP > 0.9, with the number of fine-mapped SNPs correlated strongly with the number of GWAS loci identified by LD clumping (**Supplementary Fig. 18-19**). However, 1,158 of these SNPs were not GWAS lead SNPs, and 14.9% located outside GWAS loci (1Mb regions around independently significant SNPs) (**Fig. 6a**). Consistent with our simulation results (**Fig. 3a-c**), this highlights the importance of conducting genome-wide fine-mapping to capture all relevant signals. Notably, 469 fine-mapped SNPs (25.8%) were associated with multiple traits, suggesting prevalent pleiotropy in the human genome. Moreover, the MAF of pleiotropic SNPs decreased as the number of affected traits increased (**Supplementary Fig. 20**), consistent with a model of negative selection, where highly pleiotropic variants are either purged from the population or kept at low frequencies^46^.

**Figure 6.**
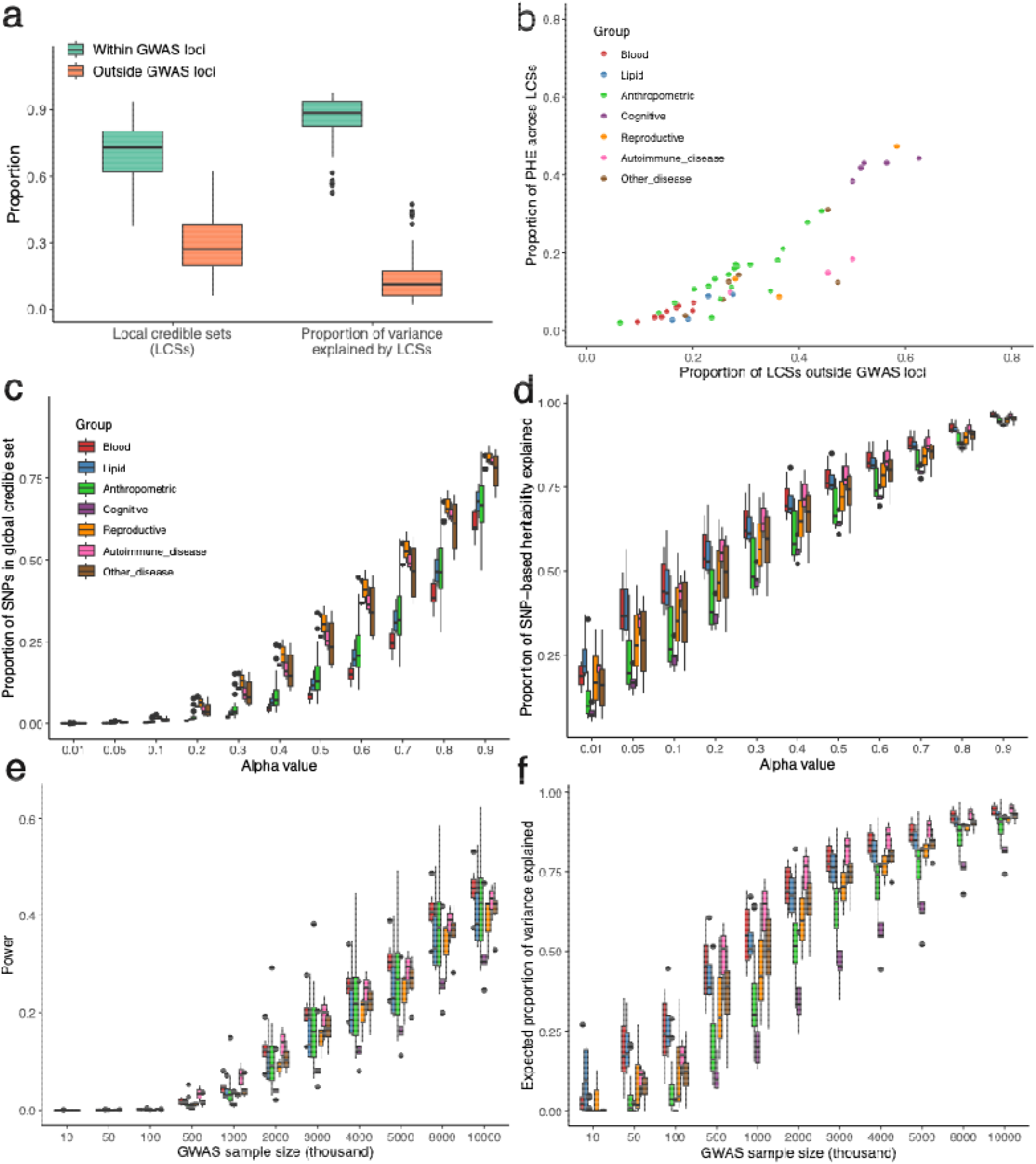
Identification of local credible set and genome-wide credible set SNPs, and theoretical prediction of fine-mapping power for 48 independent complex traits. Panel (a) shows the proportion of the identified local credible sets (LCSs) within or outside the GWAS loci (left) and the proportion of variance explained by the identified LCSs within or outside the GWAS loci (right). Panel (b) shows the relationship between the proportion of LCSs outside GWAS loci and proportion of PHE across the 48 complex traits. Panel (c) shows the proportion of identified GCS SNPs at different alpha threshold (proportion of causal variants captured) for the 48 complex traits (average sample size = 291K). Panel (d) shows the proportion of 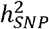 explained by the GCS SNPs at different alpha threshold. Panel (e-f) show theoretical prediction of power and proportion of SNP-based heritability explained by LCS SNPs at different GWAS sample sizes for the 48 complex traits, respectively. Colours indicate different trait categories.

We identified 19,863 0.9-LCSs with a median size of 5 SNPs, of which 29.8% were located outside the genome-wide significant loci (**Fig. 6a**). While these LCSs captured only 0.9% of causal variants, they explained 17.7% of 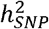, with 2.7% PHE (relative proportion of PHE = 2.7%/17.7=15.3%) attributed to LCSs in non-significant GWAS regions (**Fig. 6a**). Across trait categories, cognitive traits had the highest proportion of LCSs and PHE observed outside GWAS loci (**Fig. 6b**), consistent with the high polygenicity of cognitive traits reported previously^3^. On average, the 0.1-GCS (expected to contain 10% of all causal variants) comprised ∼1.5% of the SNPs, explaining 44.7% of the 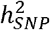 (**Fig. 6c,d**). Reproductive traits had the largest 0.1-GCSs (2.3% of SNPs for 10% causal variant coverage), while blood cell traits had the smallest (0.55%) but explained a large proportion of 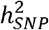 (54.6%), highlighting the impact of genetic architecture on fine-mapping uncertainty.

Our results recapitulated previous findings and led to new discoveries. For example, we identified 13 SNPs at PIP > 0.9 for SCZ from the latest meta-analysis, 5 of which overlapped with SNPs identified by FINEMAP in their study^39^, and all 8 FINEMAP-identified SNPs were included in our 0.9-LCSs. Among the 5 novel fine-mapped SNPs not identified by FINEMAP based on individual PIP, 3 were missense variants (**Supplementary Fig. 21a-d**). We also identified novel putative causal variants for CD (**Supplementary Table 4**), fine-mapping 15 SNPs, including 5 missense variants, and successfully recapitulated all 3 variants identified in a previous study using the same dataset^41^. Notably, at the NOD2 locus, we identified 4 putative causal variants (Supplementary Fig. 22), consistent with the well-established role of this locus in CD etiology. As further support for our method, we identified non-synonymous variants in 11 well-established causal genes for Crohn’s disease, schizophrenia, height, and HDL (**Supplementary Table 5**).

Leveraging the estimated genetic architecture for these 48 traits, we predicted the power of prospective fine-mapping studies. With a sample size of 2 million individuals, we predict an average fine-mapping power of 10.4% (**Fig. 6e**) and an average PHE of 57.9% (**Fig. 6f**). The predicted power and PHE varied substantially across trait categories: blood cell traits had both the highest power (12.8%) and PHE (69.4%), while cognitive traits had the lowest (5.2% power, 35.9% PHE). To achieve a PHE of 50%, blood cell traits required 1 million individuals, while cognitive traits required 4 million. For PHE = 80%, the required sample sizes increased to 3 million for blood cell traits and 8 million for cognitive traits (**Fig. 6f**).

### Incorporating functional annotations improves fine-mapping

SBayesRC incorporates functional annotations by learning their weights from the data, rather than relying on preassigned values. We observed multiple lines of evidence supporting the contribution of functional annotations to fine-mapping. First, although the impact of annotations varied across traits, certain functional annotations consistently had stronger impact on SNP effect weighting (**Supplementary Fig. 23**). Second, the 1,820 fine-mapped SNPs showed greater enrichment than GWAS significant SNPs in functional genomic regions, such as coding sequences, transcription start sites, non-synonymous variants, and conserved regions, and were more depleted in repressed regions (Fig. 7a). Third, fine-mapping power enrichment was strongly correlated with per-SNP heritability enrichment (r = 0.93), which is not subject to ascertainment bias (**Supplementary Fig. 24**). Fourth, among 13 fine-mapped SNPs for SCZ, running SBayesR (without annotations) identified 4 at PIP > 0.9 and the other 7 at lower but still meaningful PIP values (>0.5), suggesting that functional annotations enhanced fine-mapping power. Finally, some putative causal variants could not be distinguished from SNPs in high LD without considering functional annotations, as demonstrated in the examples below.

**Figure 7.**
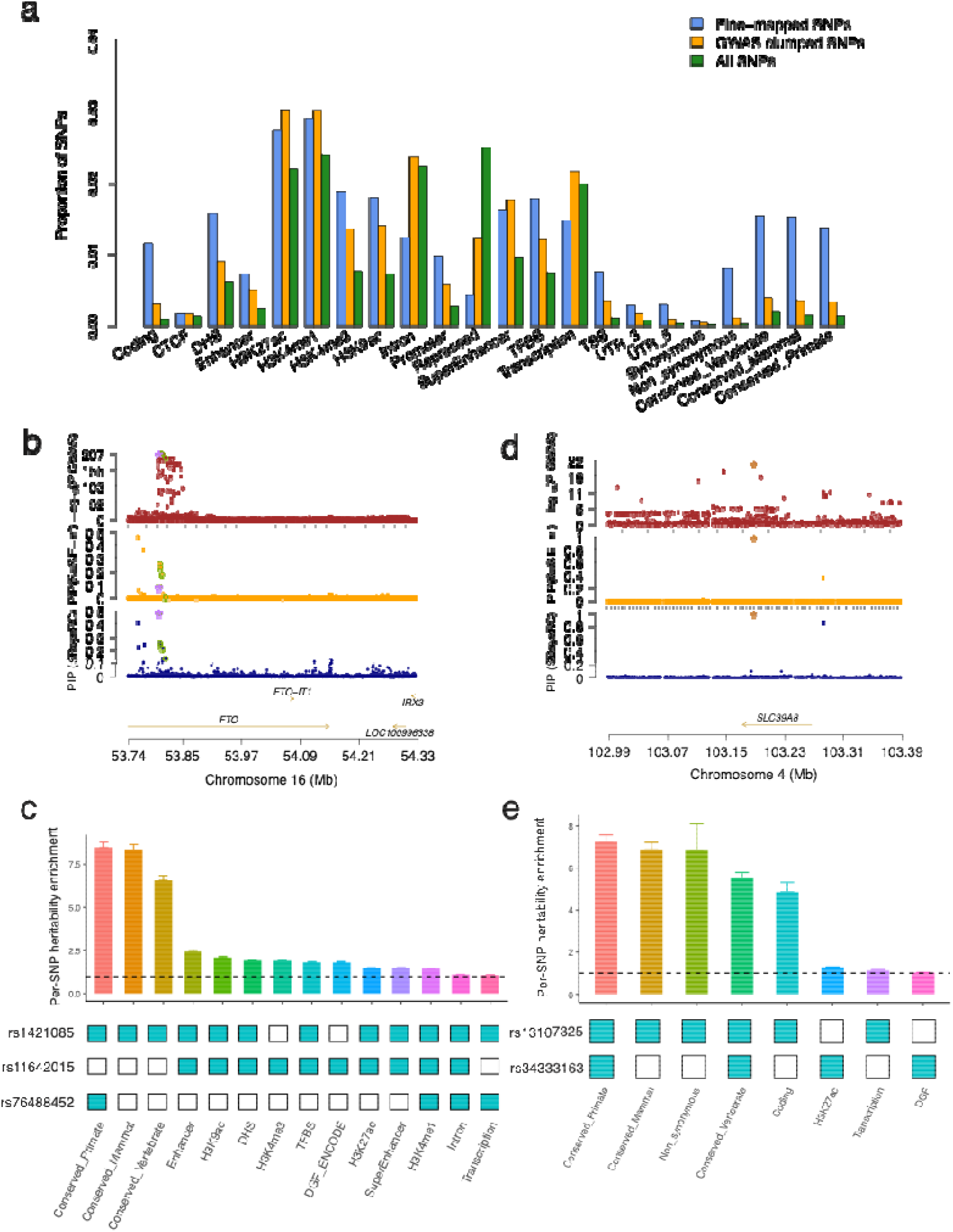
Genome-wide fine-mapping with functional annotations helped pinpoint the putative causal variants. Panel (a) shows enrichment of the genome-wide fine-mapped SNPs from SBayesRC and GWAS clumped SNPs in the 22 main functional categories defined in the LDSC baseline model. Panel (b) shows the prioritized causal variant at the *FTO* locus for BMI. The top track shows the *FTO* locus plot of the standard GWAS for BMI, the second track shows the similar plot but with the PIP from SBayesRC for BMI, and the third track shows the similar plot with PIP from SuSiE-inf. The dots with green circles are credible set SNPs identified by SBayesRC. The starred SNP is the known causal variant (rs1421085) for obesity at the *FTO* locus supported by previous functional studies. Panel (c) shows the per-SNP heritability enrichment for the causal variant (rs1421085), the GWAS lead variant (rs11642015) and the secondary signal (rs76488452) for BMI at the *FTO* locus. The annotations on the x-axis were those present at least once in these three variants, excluding quantitative annotations and duplicated annotations with flanking windows. Panel (d) shows the prioritized causal variant at the *SLC39A8* locus for SCZ. The top track shows the *SLC39A8* locus plot of the standard GWAS for SCZ, and the second track shows the similar plot but with the PIP from SBayesRC for SCZ. The starred SNP is the missense variant (rs13107325) fine-mapped for SCZ at the *SLC39A8* locus. Panel (e) shows the per-SNP heritability enrichment for the causal variant (rs13107325) and the secondary signal (rs34333163) for SCZ at the SLC39A8 locus.

One notable example is the variant rs1421085 at the *FTO* locus, a known causal variant for body mass index (BMI)^47^, which was fine-mapped in a credible set from our analysis for body fat percentage (BFP), hip circumference (HC) and waist circumference (WC). Experimental evidence supports its causal role in regulating adipocyte thermogenesis^47^. Unlike the standard GWAS results, where many SNPs in *FTO* reached genome-wide significance, both SBayesRC and SuSiE-inf identified a 5-SNP 0.9-LCS that included rs1421085 for BMI, along with four additional SNPs in almost complete LD (minimum LD *r*^2^=0.997) and with strong annotation overlaps (**Fig. 7b**). Notably, rs1421085 was prioritized in SBayesRC (PIP=0.47), likely due to its cross-species conservation annotations (**Fig. 7c**), whereas SuSiE-inf prioritized the GWAS lead SNP instead, underscoring the importance of incorporating functional annotations into fine-mapping.

Additionally, SBayesRC identified a secondary signal with another 0.9-LCS, which aligned with the result of SuSiE-inf but had not been previously reported. The lead SNP rs76488452 (PIP=0.41) resided in a conserved region across primates and was significant in COJO analysis^48^ (*P*=1.8×10^−17^) when conditioned on rs1421085. Notably, rs76488452 was only nominally significantly in GWAS marginal analysis (*P*=3.6×10^−4^), and its trait-increasing allele was in negative LD (r=-0.16) with that of rs1421085, indicating a masked effect at the SNP^49^ (estimated masked effect size *b*_2_–*r*×*b*_1_ = 0.02, consistent with the reported GWAS marginal effect size).

We highlight additional examples where functional annotations improved fine-mapping. We recapitulated a missense variant (rs13107325) in SLC39A8, a gene implicated in the latest SCZ analysis for its function in regulating dendritic spine density^50,51^. This variant was identified through an aggregated effect from multiple key functional annotations at the SNP (**Fig. 7d-e**). Another notable SCZ variant is rs11854184 in *SECISBP2L* (**Supplementary Fig. 21a**), a gene highly expressed in brain-related tissues (**Supplementary Fig. 25**). *SECISBP2L* is specifically expressed in differentiating oligodendrocytes, where the SECISBP2L-DIO2-T3 pathway regulates oligodendrocyte differentiation during myelin development^52^. SNP rs11854184 is a missense variant conserved across primates and mammals, with a high PIP (0.91) despite not reaching genome-wide significance (P-value = 1.6e-6). Although the over-expression of *SECISBP2L* in the brain can be coincidental and further experimental validation is needed to confirm causality, this finding suggests that variants below genome-wide significance can still be biologically relevant and merit fine-mapping analysis. Moreover, compared to a recent exome-wide association study for CD^53^, we identified a novel gene (*SLAMF8*) with missense putative causal variants (Supplementary Fig. 26). These results highlight the power of SBayesRC in identifying putative causal variants and provide a valuable resource for downstream analysis and functional validation.

## Discussion

In this study, we comprehensively evaluated GWFM using SBayesRC through extensive simulations and real data analyses, compared with existing fine-mapping methods that analyze one genomic region at a time. SBayesRC demonstrated superior PIP calibration, leading to better FDR control, particularly when the genetic architecture deviated from model assumptions (**Fig. 2**). Across various metrics, SBayesRC outperformed the best alternative method (SuSiE-inf) by 194% in mapping power, 21% in resolution, and 68% in precision (**Fig. 3**). In real trait analyses, SBayesRC achieved higher replication rates, lower estimation bias, and greater prediction accuracy using fine-mapped SNPs in independent validation samples (**Fig. 4**), identifying putative causal variants missed by other methods (**Fig. 7**). To date, most fine-mapping studies have focused exclusively on genome-wide significant GWAS loci^6-12^. However, our analysis revealed that 30% of the local credible sets (median size=5) identified by SBayesRC were beyond genome-wide significant loci, contributing to 15% of the 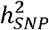 captured by all local SNP credible sets (**Fig. 6a**). These findings suggest GWFM as a more effective approach than current fine-mapping strategy that focuses solely on GWAS loci or individual regions, and highlight SBayesRC as a reliable and superior method for GWFM analysis.

The advantage of GWFM over GWAS loci-based fine-mapping arise from several key aspects. First, incorporating functional annotations improves fine-mapping power. Unlike GWAS, which does not consider functional annotations, SBayesRC incorporates functional genomic information, improving SNP effect weighting and the ability to identify causal variants that would otherwise be overlooked in GWAS. Second, fine-mapping can identify causal variants masked by LD that do not reach genome-wide significance in GWAS. Previous works^49,55^ have shown that the LD masking between causal variants is common in complex traits, which can weaken their marginal association signals. However, fine-mapping methods that assess joint association signals across SNPs can recover these masked effects, identifying causal variants with strong PIPs despite their weaker marginal GWAS signals. Third, P-value and PIP serve different purposes in managing false positives. The genome-wide significance threshold in GWAS is designed to control of false positive rate under the null hypothesis, suffering from multiple testing burdens^2^. In contrast, PIP directly controls the FDR within a Bayesian framework, automatically accounting for multiple comparisons^54^. For example, with a PIP threshold of 0.9, 90% of identified variants are expected to be true positives, while 10% may be false positives. In comparison, GWAS significance threshold is often more conservative, potentially leading to missed true causal variants. Together, we recommend directly performing GWFM analysis, rather than restricting fine-mapping to genome-wide significant loci identified from GWAS.

The advantages of SBayesRC over the region-specific fine-mapping methods is likely due to the following factors. First, SBayesRC involves a genome-wide analysis fitting all SNPs jointly across LD blocks. Compared to the existing practice focusing on 1-2 Mb regions^6-12^, using LD blocks better accounts for long-range LD, including the MHC region which is often excluded from the analysis^10,39^ (**Supplementary Fig. 27**). Although in this study all method comparisons were based on using same LD blocks as fine-mapping regions, SBayesRC benefits from genome-wide fitting which allows for better estimation of genetic architecture parameters from the full set of SNPs. Second, SBayesRC assumes a more realistic distribution of SNP effects through using MAF/LD groups along with other functional annotations. Unlike other fine-mapping methods^6-12^, which either ignore annotation data or lack a unified framework to integrate it with GWAS data, SBayesRC estimates the impact of functional annotations and SNP effects within the same model, enabling a proper Bayesian learning process. This framework estimates, instead of preassign, weights to trait-relevant annotations, and is robust to the inclusion of irrelevant annotations (**Fig. 2a**). Third, SBayesRC utilizes Gibbs sampling algorithm to estimate model parameters and PIPs, which is asymptotically exact. In contrast, FINEMAP and FINEMAP-inf use shotgun stochastic search, while both SuSiE and SuSiE-inf rely on variational Bayes. Previous studies have demonstrated that MCMC sampling generally leads to a higher accuracy in capturing the posterior distribution than variational Bayes^27^.

In addition to the improved fine-mapping performance, we introduced new features for GWFM using MCMC sampling. First, based on the posterior samples of SNP effects, we refined LCS by PEP, ensuring that all included SNPs explain more 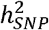 than a random set. This approach is similar but more interpretable than the “purity” parameter in SuSiE (**Supplementary Note 1; Supplementary Fig. 28-29**). Second, while a LCS quantifies local uncertainty in identifying a causal variant, a GCS quantifies uncertainty in identifying all causal variants, reflecting both the power of current study and the genetic architecture of the trait. However, computing GCS requires an estimate of the total number of causal variants, which is better achieved when analysing all SNPs jointly in the model. Third, similar to estimating genome-wide polygenicity and 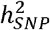, we estimated the proportion of polygenicity and 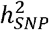 explained by LCSs and GCSs. The analysis of 48 complex traits showed that although 19,863 0.9-LCS were identified, they captured only 1% of all common causal variants, explaining 18% genetic variance, indicating that many causal variants with small effects remain undiscovered. Fourth, the genetic architecture estimation enables power calculations for fine-mapping, informing the sample size required to identify a certain proportion of causal variants or, 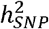 explained in future studies. The robustness of this approach is supported by real data projections aligning with theoretical predictions (**Fig. 5**). Finally, to address poor mixing in standard Gibbs sampling when analyzing SNPs with extremely high LD, we implemented a scalable tempered Gibbs sampling algorithm^32^ and assessed the convergence of each SNP’s PIP using multiple independent chains, facilitating robust posterior inference.

We note several limitations of this work. First, there are certainly more complicated scenarios about effect size distribution for causal variants that have not been investigated in our simulations. However, SBayesRC remains one of the most flexible models available, as its assumed multi-component Gaussian mixture is well-suited to accommodate various scenarios. Second, unlike individual-level models where each PIP is estimated conditional on all other SNPs jointly, SBayesRC is a summary-level model that ignores LD between LD blocks. As a result, SNPs outside the block influence PIP estimates only through the mixture distribution of SNP effects. Third, we applied SBayesRC only to GWAS summary data from relatively homogenous populations (inferred European ancestry) and the robustness of the methods on GWAS data based on trans-ancestry meta-analyses is not investigated. Future work should evaluate its performance in diverse populations. Fourth, SBayesRC requires LD information from a reference cohort that matches with GWAS ancestry without substantial sampling variation. Our simulation showed that when using a LD reference of thousands of samples (n=3,642), SBayesRC remained robust (**Supplementary Fig. 30**). However, with a smaller LD reference (n=500), we observed some inflation in PIPs, indicating the importance of sufficient LD reference sample size for maintaining accuracy. Fifth, to construct LCS, a LD threshold was arbitrarily chosen to define a set of SNPs in LD with the candidate causal variant. Although we have explored the robustness to this threshold (**Supplementary Fig. 3**), future methodological advancements, such as Bayesian hypothesis testing with hierarchical clustering^56^, could be used to relax this condition. Sixth, our power prediction is based on the genetic architecture estimates given a SNP set. However, as sample sizes increases, more common SNPs may be detected, potentially affecting polygenicity and SNP-based heritability estimates. Seventh, although we incorporated a comprehensive set of functional annotations from BaselineLD (v2.2)^57^, these annotations remain incomplete in genome coverage and lack tissue and cell-type diversity. Incorporation of context-dependent annotations, such as those derived from single-cell data, could further enhance fine-mapping accuracy by capturing cell-type-specific regulatory effects. Despite these limitations, our study provides a powerful and robust GWFM framework for identifying causal variants, highlighting the advantages of this approach over existing region-specific fine-mapping methods. With its capacity to enhance mapping power in the current study and to predict mapping power for future studies, we anticipate that GWFM using a state-of-the-art GBMM will become the preferred method for fine-mapping complex traits.

## Methods

### Ethics approval

The University of Queensland Human Research Ethics Committee B (2011001173) provides approval for analysis of human genetic data used in this study on the high-performance cluster of the University of Queensland.

### Low-rank GBMM

We used state-of-the-art GBMM that employed a low-rank model to improve computational efficiency and robustness^24^. As described below, the low-rank GBMM can be derived from the individual-level model. Consider a multiple linear regression of phenotypes on genotypes:

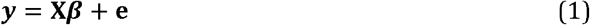

where **y** is an *n* × 1 vector of complex trait phenotypes and **X** is an *m* × *n* matrix of mean-centred genotypes, **β** is *m* × 1 vector of SNP effects on the trait, and **e** is *n* × 1 vector of residuals with 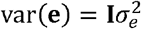. Let

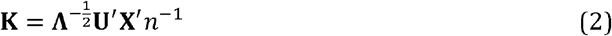

where **Λ** and **U** are diagonal matrix of eigenvalues and matrix of eigenvectors for the LD correlation matrix **R = X ′X**/*n*^1^, respectively. It follows that **K ′K= P***n* ^−1^, where **P** is the projection matrix of **y** on the column space of **X**, and **KK = I***n*−. Multiplying both sides of Eq (2) by **K** gives

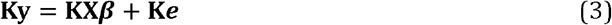

or

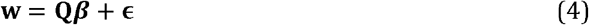

When only the top *q* principal components of **R** are used, the dimension of **w** is *q*×1 and **Q s**is *q*×*m*. By selecting *q* ≪ *n*, this model would have a substantially lower rank than Eq (1), improving the computational efficiency for the estimation of ***β***. By default, our method automatically determines *q* through pseudo cross-validation based on GWAS summary statistics(Supplementary Note 10 of Zheng et al. ^24^). With a recognition that **b = X′y**/*n* is the GWAS marginal effect estimates, **w** can be directly computed from the GWAS summary statistics

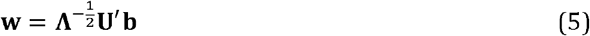

with **Λ** and **U** obtained from a LD reference sample, and can be computed as

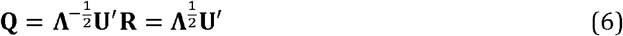

Essentially, the low-rank model transforms the mutually correlated GWAS marginal effects (**b**) into a set of independent data points (**w**), while maintaining the ability to estimate individual SNP joint effects (**β**). In practice, we compute **w** and **Q** within quasi-independent LD blocks in the human genome. With this low-rank model, we can estimate ***β*** for all common variants jointly through considering ***β*** as random effects. In addition, this model allows a direction estimation of the residual variance, as 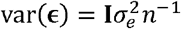, which can be used as a nuisance parameter to accommodate heterogeneity in the summary statistics and LD reference^24^.

### SBayesC, SBayesR, and SBayesRC

GBMMs are flexible in specifying the prior distribution of SNP effects. In SBayesC, the prior for the effect of SNP *j* is given by

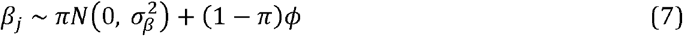

where 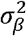 is the common variance across all causal variants, π is the proportion of SNPs with nonzero effects, and *ϕ* is a point mass at zero. Both 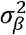 and *π* are considered as unknown, with 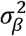 following a scaled inverse chi-squared prior distribution and *π* following a uniform prior distribution^24^. This is also the underlying model for most fine-mapping methods, assuming that only a fraction of SNPs are causal with nonzero effects.

SBayesR^22^ extends SBayesC by assuming a more flexible prior distribution for SNP effects, using a multi-component Gaussian mixture:

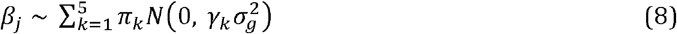

where 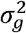 is the total genetic variance, *γ* = [0,10^−5,10−4,10−3,10−2^]’ are pre-specified scale factors that constrain the variance of each effect size distribution, representing zero, very small, small, medium, and large effect size categories, and *π*_*k*_ is the prior probability of SNP *j* belonging to the *k*th distribution. With different *π*_*k*_ values, this multi-component mixture model can approximate almost any distribution of SNP effects. This allows SBayesR to better model genetic architectures with a wide range of effect sizes compared to SBayesC.

To further account for the stratification of causal variants and their effects based on functional annotations, SBayesRC^24^ assumes a SNP-specific prior probability of distribution membership, *π*_*jk*_, which depends on the annotations at each SNP through a generalised linear model:

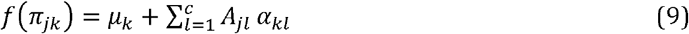

where *f* (·) is the probit link function, *μ*_*k*_ is the intercept, *A*_*jl*_ is the value of annotation on SNP (either binary or continuous annotations), and *α*_*kl*_ is the effect of annotation *l* on the prior probability of SNP *j* belonging to the *k*th distribution. To enable a data-driven estimation of annotation effects, we assume an independent and identical normal prior for each annotation effect,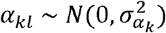. This prior assumes, a *priori*, an equal contribution of all functional annotations to the SNP prior probability of membership. Details of the method and the MCMC sampling scheme can be found in ref^24^.

In this study, we ran SBayesRC with 4 MCMC chains, each consisting of 3,000 iterations with the first 1,000 discarded as burn-in. Analysing 13 million SNPs with 96 annotations, SBayesRC required only 150 GB of RAM and 13 hours of computation using 24 CPU cores, which are commonly available in a standard computing cluster.

### LD blocks

We partitioned the genome into 1,588 approximately independent LD blocks, following the methods of Li et al^13^ and Berisa and Pickrell^58^. Specifically, we merged the LD blocks from Li et al. to ensure a minimum length of 1 Mb and treated the MHC region as a single LD block due to its complex LD patterns, which had the largest number of SNPs. As a result, our LD blocks ranged in size from 1 to 30 Mb, with a median length of 1.5 Mb.

### Calculation of PIP

We assessed the strength of joint association of each SNP using PIP, i.e., the probability of a SNP being included with a nonzero effect in the model, given the data (**w**). Let *δ*_*j*_ be the indicator variable for the distribution membership for SNP *j* with*δ*_*j*_ = 1 indicating a null effect and *δ*_*j*_ = 2,…, 5 indicating a nonzero component. In SBayesRC, we computed PIP for SNP *j* as

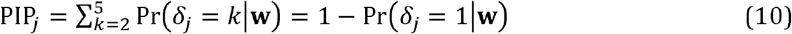

In the literature, Pr (*δ*_*j*_ = 1| *Data*), the probability of SNP _*j*_ having a zero effect, is often computed by counting the frequency of *δ*_*j*_ = 1 across MCMC samples^23,25^. To improve precision, we use Rao-Blackwellized estimates^59,60^ and compute the posterior mean of [1 − pr (*δ*_*j*_ = 1|**w, *θ*)]** conditional on data and all the other parameters except *β*_*j*_ (***θ*)** across *T* iterations. That is, suppose 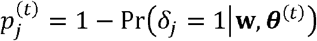, we have

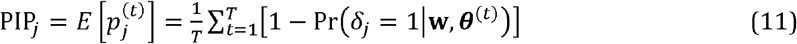

where

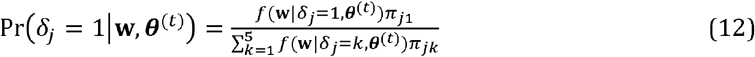

with *f*(**w**| ***δ***_*j*_ = k, ***θ***^**(*t*)**^ being the likelihood given *δ*_*j*_ and the sampled values of other parameters. A detailed derivation can be found in **Supplementary Note 5**.

To further improve robustness, we run multiple MCMC chains simultaneously, randomly shuffling the sampling order of SNP effects in every iteration. For SNP *j* in chain *s* at iteration *t*, we have

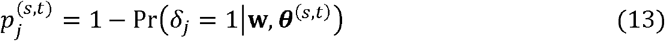

Following Gelman and Rubin (1992)^61^, we calculated the potential scale reduction factor (PSRF) for each SNP PIP:

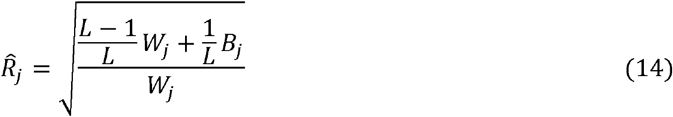

where 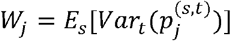 is the averaged within-chain variance across chains, and 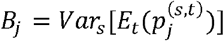 is the variance of the means of the chains. By comparing the between-chain variance to the within-chain variance, PSRF assesses whether the MCMC chains have converged to a stationary distribution. Empirically, a PSRF value below 1.2 indicates adequate convergence^61^. By pooling samples across chains, our final PIP estimate is

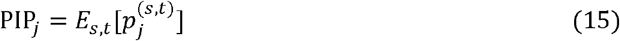

We report the PSRF value to imply if the SNP PIP is converged or not. SNPs with high PIP but large PSRF should be interpreted with caution. Running longer chains is recommended if a remarkable proportion of fine-mapped variants have high PSRG values. Across 48 well-powered complex traits analyzed, we found that none of fine-mapped individual SNPs (singleton LCSs) had PSRF < 1.2, while 99.4% of SNPs included in the local credible sets had PSRF < 1.2 and 99.9% had PSRF < 2, suggesting adequate convergence in our analyses.

### Heuristic estimation of mixture components

The standard SBayesRC requires pre-specifying the number of mixture components before analysis. While polygenic prediction is generally robust to this choice^24^, fine-mapping may be affected if an unnecessary small-effect component is included, leading to null SNPs being misclassified and inflating PIP. This could bias the estimated number of causal variants. To address this, we implemented a heuristic procedure for determining the number of mixture components in SBayesRC. Multiple models, varying from 2 to 5 mixture components, were run in parallel for 500 MCMC iterations. The posterior distribution of 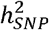 was then computed to assess model fit. The model with the highest posterior mean, where the lower bound (posterior mean minus standard error) was greater than that of the second-best model, was selected for formal analysis. If no significant difference was found, the model with the fewest components was chosen.

### Tempered Gibbs sampling

Joint analysis of all common variants presents a challenge for MCMC mixing when using the standard Gibbs sampling (GS) algorithm. For example, in a finite number of iterations, a common causal variant may fail to enter the model if the SNP in complete LD with it has already been selected. To address this issue, we incorporated a tempered Gibbs sampling (TGS) algorithm^32^ into our SBayesRC model. Essentially, TGS improves mixing by (1) strategically selecting SNPs for updating (“informed choice”) – focusing on those whose indicator variable *δ*_*j*_ is currently in the tail of the full conditional distribution, and (2) allowing for longer jumps across local maxima by sampling *δ*_*j*_ from a tempered distribution. For computational efficiency, we applied TGS only to SNPs with nonzero effects sampled from the standard GS to assess whether alternative SNPs in high LD (r^2^>0.95) could provide a better fit. Further details on our TGS implementation and the corresponding pseudocode are provided in Supplementary Note 6.

### Local credible sets

Following prior work^9^, we defined LCS at confidence level *α* (*α*-LCS) as the minimum set of SNPs that contains at least one causal variant with probability *α*. To construct an *α*-LCS, SNPs were ranked by their PIPs. For a focal SNP that has the highest PIP and not yet assigned to an LCS, a candidate set was formed by including all remaining SNPs in high LD (r^2^ > 0.5) with the focal SNP. The *α*-LCS was then determined by summing PIPs in decreasing order until the total reached at least α. This process was iterated over SNPs until no possible LCS can be formed. For each *α*-LCS, we computed a posterior SNP-heritability enrichment probability (PEP) that the LCS explains more 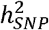 than a random set of SNPs with the same size. We reported all 0.9-LCSs with PEP > 0.7 for each LD block. A schematic of the procedure is shown in **Supplementary Fig. 1**, differences to existing approaches are discussed in **Supplementary Note 1**, and further justification of these thresholds based on simulations is provided in **Supplementary Note 2**.

### Global credible sets

The *α*-GCS identifies the smallest set of SNPs that capture *α*% of the causal variants, quantifying the overall uncertainty in fine-mapping and reflecting the genetic architecture of the trait. The size of *α*-GCS decreases with increasing power and a smaller fraction of small-effect variants. We computed the *α*-GCS as the cumulative sum of decreasingly ranked PIPs until the total exceeded *α* × *m*_*c*_, where *m*_*c*_ was the estimated number of causal variants from GBMM for the trait. Since each SNP is causal with probability PIP_*j*_, the expected number of causal variants in a set Ω is E [Number of causal variants in Ω] ∑_*j*∈Ω_ PIP_*j*_. Since the *α*-GCS is constructed such that ∑_*j*∈ *GCS*_ PIP_*j*_ ≥ *α* ×*m*_*c*_., this directly implies that E[Number of causal variants in *α*-GCS] ≥ *α* ×*m*_*c*_.

### Estimation of power and variance explained given the data

For the identified SNPs using individual PIP or LCS, we estimated the true positive rate (TPR or power) of identifying the causal variants at a given threshold *α*,

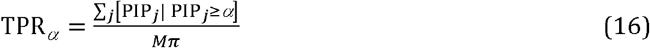

where *M* is the total number of SNPs, and *π* is the proportion of causal SNPs. A formal derivation is given in **Supplementary Note 7**.

The proportion of SNP-based heritability explained (PHE) by LCSs or GCS is estimated based on the MCMC samples of SNP effects. For a focal set (Ω) of SNPs in iteration t, we computed PHE as

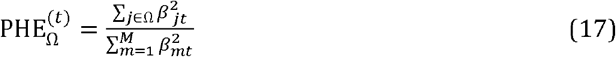

where *β*._*t*_ is the sampled effect for a SNP in iteration *t* in scaled genotype units. Finally, we computed the posterior mean across MCMC iterations as the estimate for PHE_Ω_

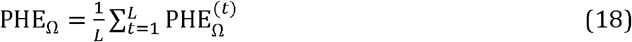

where *L* is the total number of MCMC iterations.

### Prediction of power and variance explained for prospective studies

We aim to predict the power of identifying a certain proportion of the causal variants in a prospective fine-mapping analysis, given a sample size (n) and the genetic architecture of the trait, when PIP from a GBMM is used as the test statistic. As shown in **Supplementary Note 8**, assuming that variance explained by the causal variant is *ν*, the sampling distribution of PIP from the multi-component mixture model, e.g., SBayesRC, is

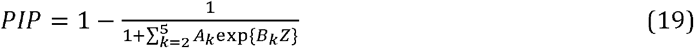

where 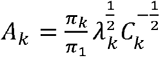 and 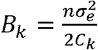 are two constants given the genetic architecture parameters(***π*** and 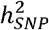), with 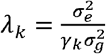 and *C*_*k*_ = *n* + *λ*_*k*_, and *Z* is a data-dependent random variable following a non-central Chi-square distribution with the non-centrality parameter 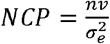.

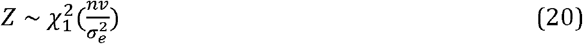

Given the threshold of PIP being *α* for the hypothesis test, the power to detect this causal variant can be calculated as

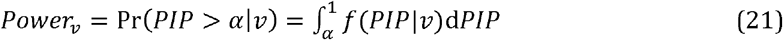

where *f* (*PIP*|*ν*) is Eq (19) above. To compute the power for identifying any causal variant, we integrated out *ν* by

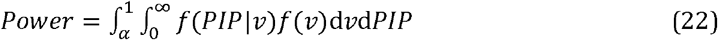

Where

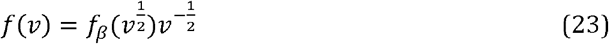

and *f*_*β*_(·) is the distribution of estimated from the SBayesRC model.

Therefore, given a sample size, the expected number of causal variants identified from fine-mapping is

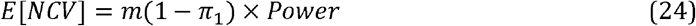

The expected proportion of genetic variance explained by the fine-mapped variants is

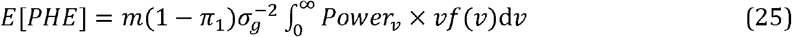

Since it is computationally challenging to obtain an analytical solution, we opted to estimate these quantities through Monte Carlo simulation (**Supplementary Note 8**). While our theoretical prediction does not model LD between SNPs, the extent to which the observed values were consistent with the predicted suggests that LD had been effectively, albeit not perfectly, accounted for by our LCSs.

### Disease sample size at the liability scale

For diseases or binary traits, we converted the GWAS summary statistics from the linear mixed model to the liability scale prior to running GBMM. Based on the method in Yang et al.^43^, we estimated the sample size at the liability scale that gives an equivalent power to detect a locus affecting a quantitative trait with the same properties,

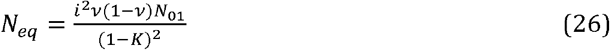

where *K* is the disease prevalence, *ν* is the sample prevalence, *i* = *h*/*K* with *h* being the height of standard normal curve at the truncation point *t* = 1 − *K*, and *N*_01_ is the total number of cases and controls. Given the z-score (*z*_*j*_) from the original GWAS summary data for SNP *j*, the marginal effect and its standard error at the liability scale can be estimated as following ref^62^

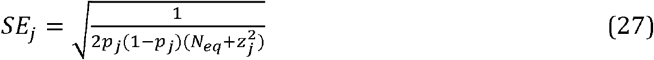

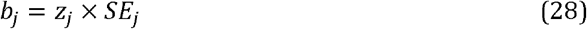

where *p*_*j*_ is the minor allele frequency of the SNP.

The results from GBMM using the converted summary statistics will be directly comparable across traits regardless of the sample prevalence and the type of traits. In our prediction analysis of power, we compared results between diseases and quantitative traits based on the equivalent sample size estimated from Eq (26). Similarly, to estimate the number of cases required, in a case-control study with equivalent number of controls, to achieve a certain power, we rearranged the same equation so that

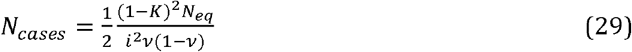

### Simulations based on imputed genotype data from the UK Biobank

To evaluate the performance of GBMM, we ran simulations using imputed genotype data from the UK Biobank after quality control (QC). We selected 300,000 unrelated individuals and included ∼1.2 million HapMap3 SNPs with MAF > 0.01, Hardy-Weinberg equilibrium test *P* > 1×10^−6^, genotyping rate > 0.95, and imputation information score > 0.8 for simulations.

We randomly *m*_*c*_ = 10.000 sampled casual variants from the genome for 100,000 individuals and simulated complex trait phenotypes based on the following model:

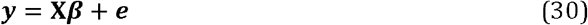

where **X** is the genotype matrix for the causal variants, ***β*** is the vector of causal variant effects, and ∼ *N*(0,**I***var*(**X*β***) /(1/*h*^2^ − 1)) with *h*_2_ = 0.5 being the proportion of phenotypic variance explained by all the causal variants. Based on this underlying model, we simulated three genetic architectures: sparse architecture, large-effects architecture, and LD-MAF stratified (LDMS) architecture. In simulations under the sparse genetic architecture, causal variant effects were drawn from *β*_*i*_∼*N*(0,*h*^2^ /*m*_*c*_), while the remaining SNPs had zero effect. For the large-effects genetic architecture, effect sizes for causal variants were drawn from two distributions, 10 random causal variant with effects from *N*(0,0.2 *h*^2^ /10) and the remaining causal variants with effects from *N*(0,0.8 *h*^2^ /9990). For the LDMS model, we partitioned all genome-wide SNPs into four LD and MAF groups (by their median values) and only sampled the causal variants from the high LD and high MAF group with the same normal distribution as in the sparse genetic architecture.

We ran a standard GWAS using the genotypes and the simulated phenotypes under different genetic architectures. We then used the GWAS summary data to perform GBMM (SBayesRC^24^, SBayesR^22^, and SBayesC) implemented in GCTB, along with SuSiE^6^, FINEMAP^7^, SuSiE-inf^8^, FINEMAP-inf^8^ and PolyFun+SuSiE^10^, to compute PIPs for detecting causal variants and estimate their effect sizes. For all methods, we followed the recommended default parameter settings. Specifically, for SuSiE and SuSiE-inf, we set the purity parameter to 0.5, the maximum number of SNPs per credible set (n_purity) to 100, and max number of causal variants to 10. We used imputed genotypes from 10,000 random samples of European ancestry from the UKB as the LD reference. To ensure a fair comparison, we used the same LD reference sample and the same independent LD blocks to define fine-mapping regions for all fine-mapping methods, irrespective of p-value significance. We repeated the whole process 100 times and quantified the true discovery rate (TDR), power, mapping precision, and replication rate for each method.

TDR was quantified as the proportion of identified *α*-LCS containing at least one causal variant, while power was defined as the proportion of simulated causal variants included in the identified *α*-LCS. Mapping precision was computed as the physical distance between the identified SNPs and nearest causal variants. The replication rate was the proportion of variants with PIP > 0.9 in a GWAS sample that were identified in an independent replication sample at the same or a lower PIP threshold.

### Real data analysis

We analysed 597 complex traits from UK Biobank using GWAS summary data from Neale’s lab (**Data Availability**) and schizophrenia^39^ and Crohn’s disease^41^ using previously published GWAS summary data. The 597 UKB traits were selected based on z-score > 4 and high-confidence heritability estimates using LD score regression^57^. We used annotations from the baseline model BaseLineLD v2.2^57^ and extract imputed SNPs with MAF > 0.001 and that overlapped with the annotations, resulting in 13,065,104 imputed SNPs passed quality control. We used 10,000 random samples from the UKB as the LD reference to run the SBayesRC and other region-specific fine-mapping analysis. Additionally, we selected 48 well-powered traits with relatively large sample size (n > 100, 000), high heritability estimate (*h*2 > 0.1), and at least one fine-mapped SNP with PIP > 0.9.

For the polygenic score prediction analysis using fine-mapped variants only, we performed quality control on the imputed genotype data provided by the UKB analysis team^30^. Following ref^24^, we kept SNPs with MAF > 0.01, Hardy-Weinberg Equilibrium test P > 10^−10^, imputation info score > 0.6 within each ancestry samples. We removed samples with mismatched sex information, samples withdrawn from participation, and cryptic related samples. The final UKB dataset consisted of 4 ancestries: European (EUR, N= 347,800), East Asian (EAS, N=2,252), South Asian (SAS, N=9,436) and African (AFR, N=7,006). For continuous traits, the phenotypes were filtered within the range of mean +/-7SD, followed by rank-based inverse-normal transformed within each ancestry and sex group. GWAS were performed using PLINK2^63^ with sex, age, and the first 10 principal component as covariates. Linear regression was used for continuous traits, while logistic regression was applied for binary traits.

## Supplementary Information

Supplementary data include 29 supplementary figures, 4 supplementary tables and 8 supplementary notes.

## Data Availability

Our SBayesRC-enabled genome-wide fine-mapping results for 599 complex traits are available at link (https://sbayes.pctgplots.cloud.edu.au/data/SBayesRC/share/Finemap/v1.1/). The UK Biobank data are available through formal application to the UK Biobank (http://www.ukbiobank.ac.uk). The GWAS summary data for 597 complex traits in UK Biobank are from http://www.nealelab.is/uk-biobank/. The LD data used in this study are available at https://cnsgenomics.com/software/gctb/#Download. All the other datasets used in this study are available in the public domain.

## Code Availability

Summary-data-based genome-wide Bayesian mixture models are implemented in a public available software GCTB at https://cnsgenomics.com/software/gctb/#Download. Methods to compute LCS and GCS have also been implemented in GCTB (https://cnsgenomics.com/software/gctb/#Genome-wideFine-mappinganalysis). Online tool for predicting fine-mapping power: https://sbayes.pctgplots.cloud.edu.au/shiny/power/.

## Acknowledgements

This research was supported by the Fundamental Research Funds for the Central Universities, the 1·3·5 project for disciplines of excellence, West China Hospital, Sichuan University (ZYYC24006, ZYJC20002), Australian National Health and Medical Research Council (1177268, 1113400,1173790), the Australian Research Council (FL180100072, DP220101947, DP230101352) and the National Institute of Mental Health (5R01MH121545-05). This study makes use of data from the UK Biobank (project ID: 12505).

## Author Contributions

J.Z. conceived and supervised the study. J.Z. and Y.W. developed the methods. J.Z., P.M.V. and Y.W. designed the experiment. Y.W. conducted all analyses with the assistance or guidance from J.Z., Z.Z., L.T., T.L., Q.F., H.C., L.Y., P.M.V., N.R.W. and M.E.G.. Y.W., and J.Z. wrote the manuscript with the participation of all authors. All the authors approved the final version of the manuscript.

