## Supplementary Materials for "Genome-wide fine-mapping improves identification of causal variants"

- Wu *et al.*

**Contents**

**Supplementary Figure 1-30**

**Supplementary Table 1-2**

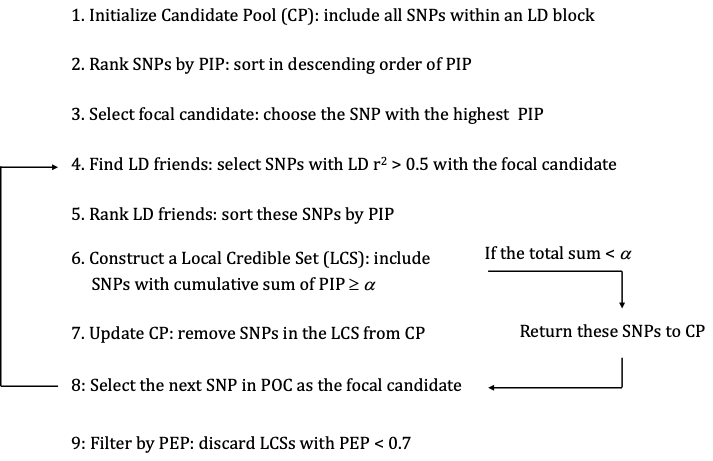

**Supplementary Figure 1** Procedure of constructing local credible sets (LCSs) using PIPs from SBayesRC.

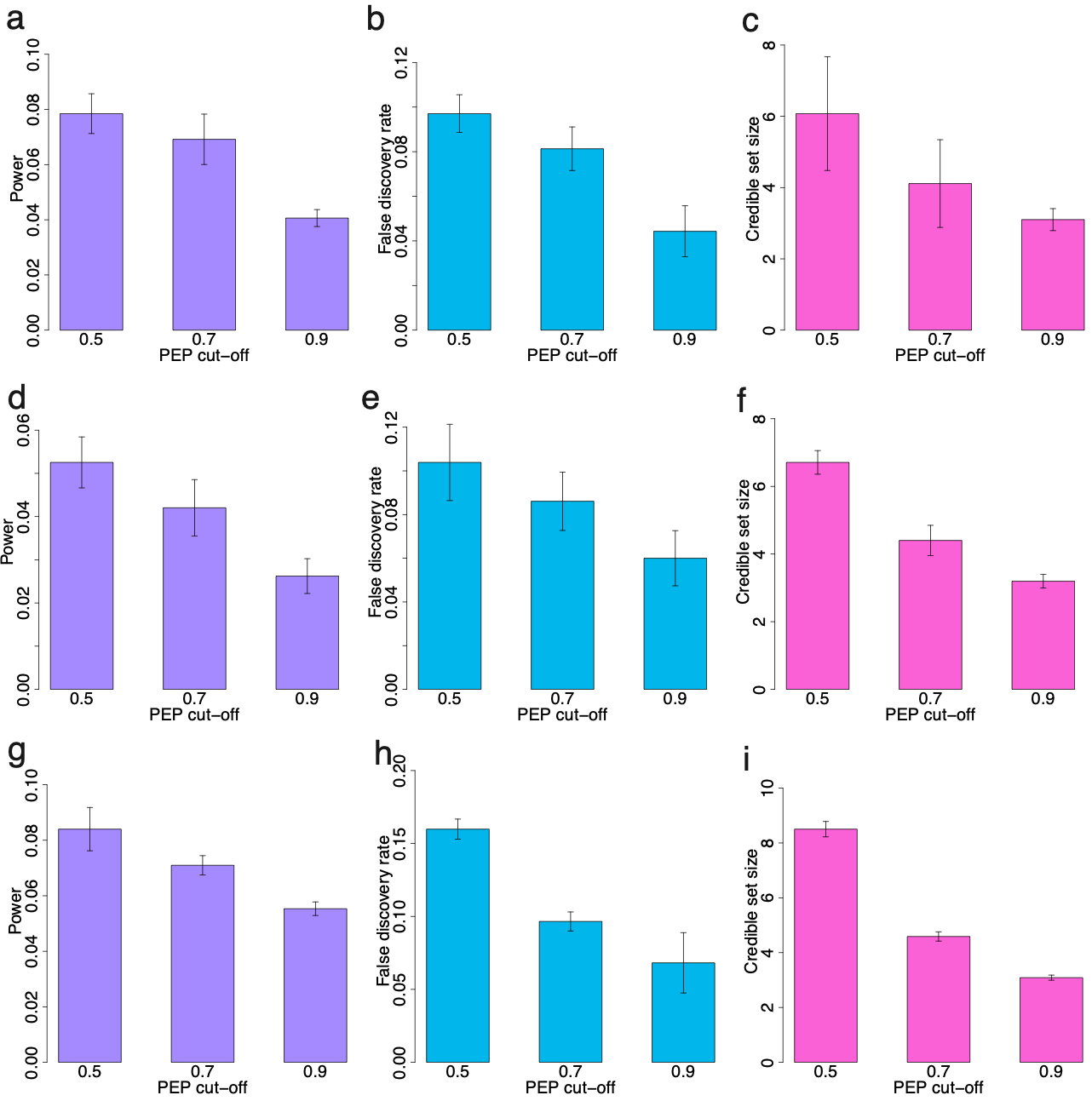

**Supplementary Figure 2** Comparison of power, false discovery rate (FDR), and credible set size (CSS) across different PEP cutoffs. Panels (a, d, g) show power, (b, e, h) show FDR, and (c, f, i) show CSS. Each row corresponds to a different genetic architecture in simulations: sparse genetic architecture (top row), large-effect genetic architecture (middle row), and LDMS genetic architecture (bottom row).

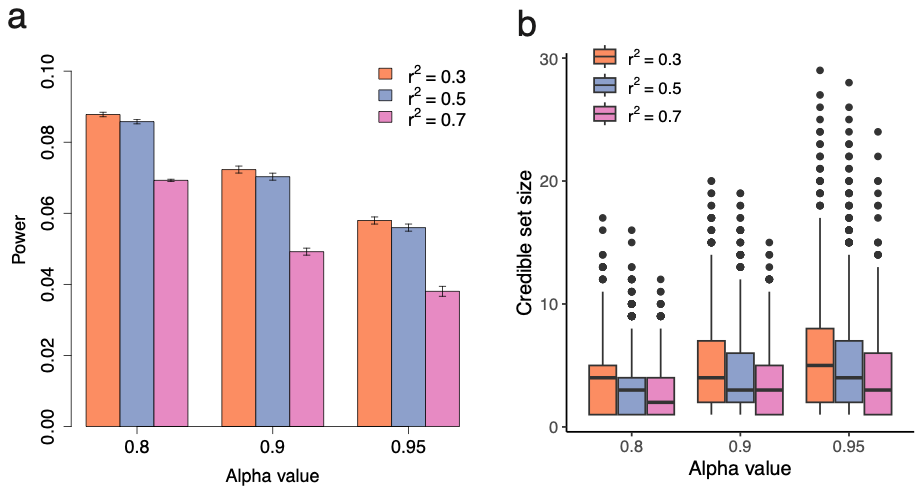

**Supplementary Figure 3** Comparison of power (a) and credible set size (b) at different LD r^2^ threshold in our GWFM using SBayesRC in simulations with sparge genetic architecture.

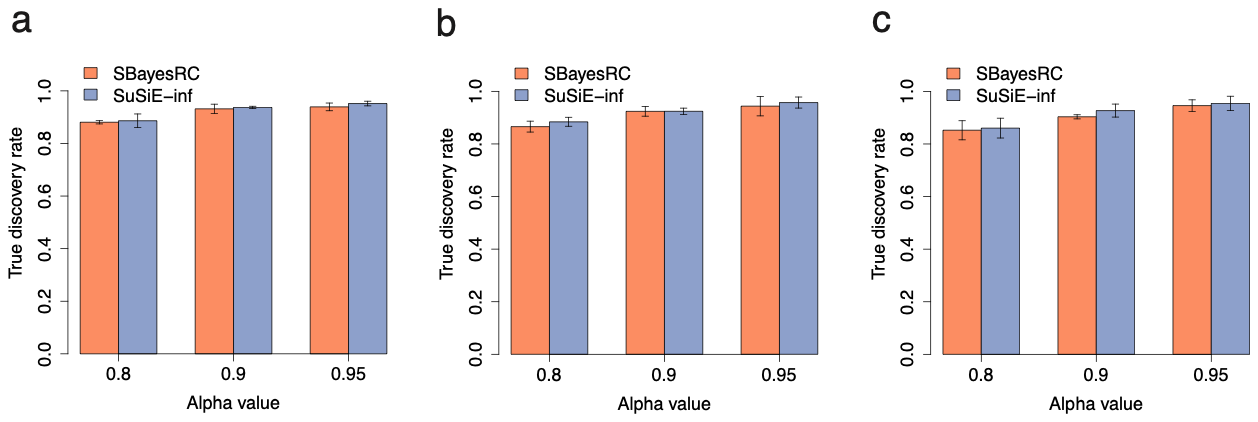

**Supplementary Figure 4** Comparison of true discovery rate in local credible set (LCS) detection between SBayesRC and SuSiE-inf. Shown in panels (a-c) are true discovery rate comparison between SBayesRC and SuSiE-inf at the same alpha cutoff. Results showed in each column correspond to the simulation under sparse genetic architecture (a), large-effects genetic architecture (b) and LDMS architecture (c).

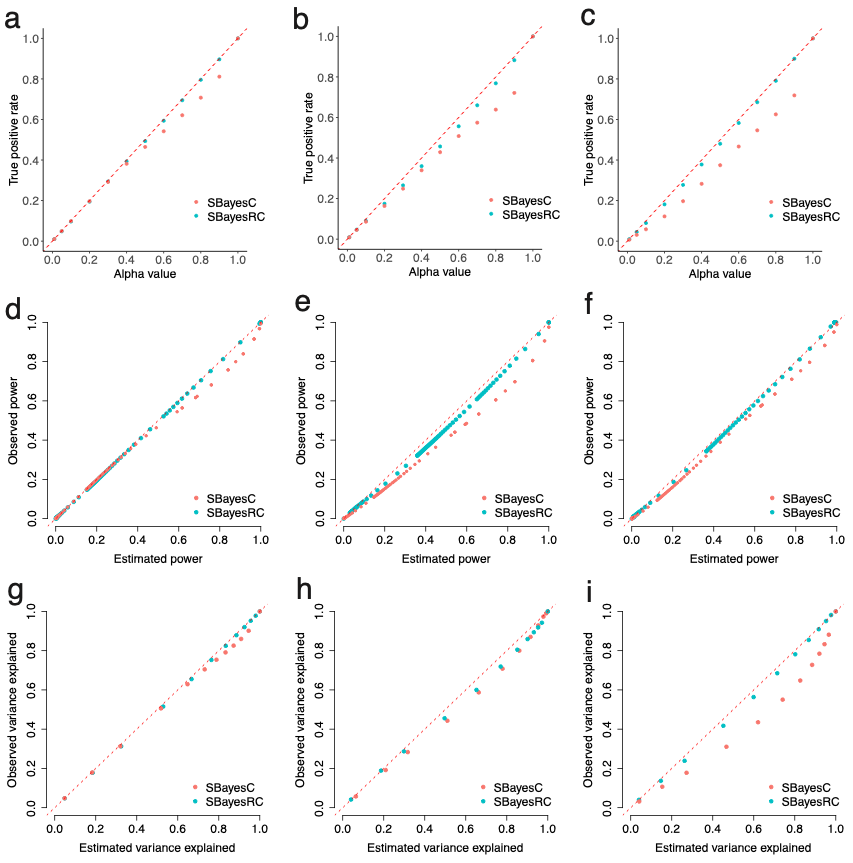

**Supplementary Figure 5** Genome-wide credible set calibration of GBMM under simulations with various genetic architectures. Panels (a-c) show the relationship between true positive rate and credible set cut off value (i.e., alpha). Panels (d-f) further show the relationship between the estimated power and observed power at a wide range of PIP cutoff. Panels (g-i) show the relationship between estimated variance explained from GBMM and observed variance explained. Results showed in each column correspond to simulation under sparse genetic architecture (a, d and g), large-effects genetic architecture (b, e and h) and LDMS architecture (c, f and i), respectively.

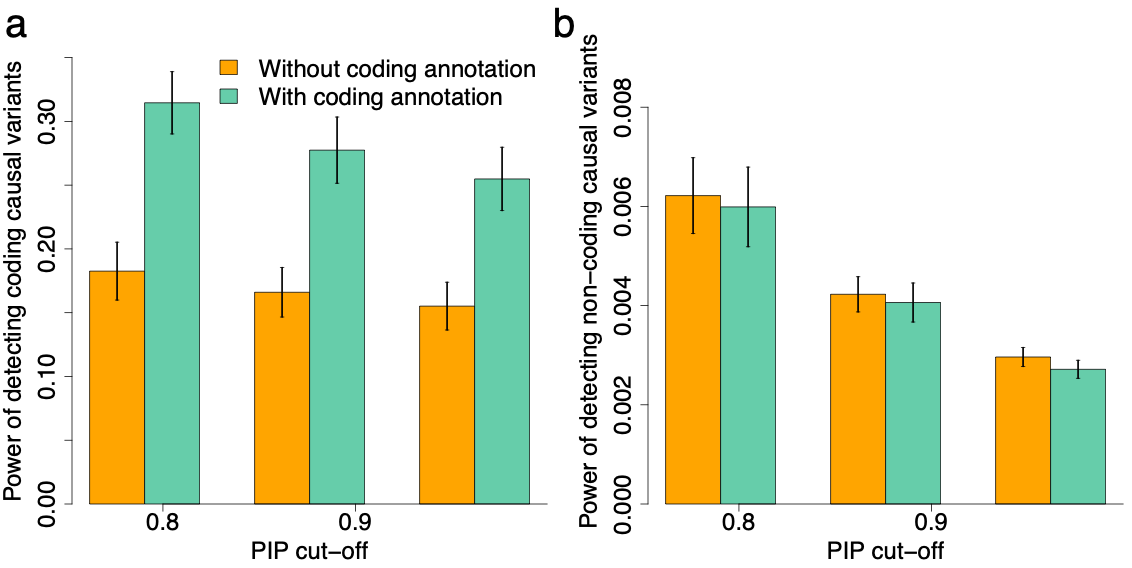

**Supplementary Figure 6** Power of detecting coding (a) and non-coding (b) causal variants with and without the coding annotation using SBayesRC at different PIP cut-off. In the simulation, 30% of the total genetic variance was allocated to the coding regions, which covered ~3.4% of the SNPs, leading to 10x enrichment in per-SNP heritability. The remaining 70% of the total genetic variance was from the noncoding regions.

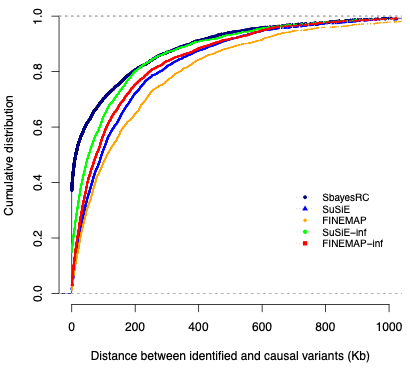

**Supplementary Figure 7** Distance between the causal variants and the SNPs identified by different methods at PIP of 0.9 in simulations with removing 50% of causal variants under sparse genetic architecture.

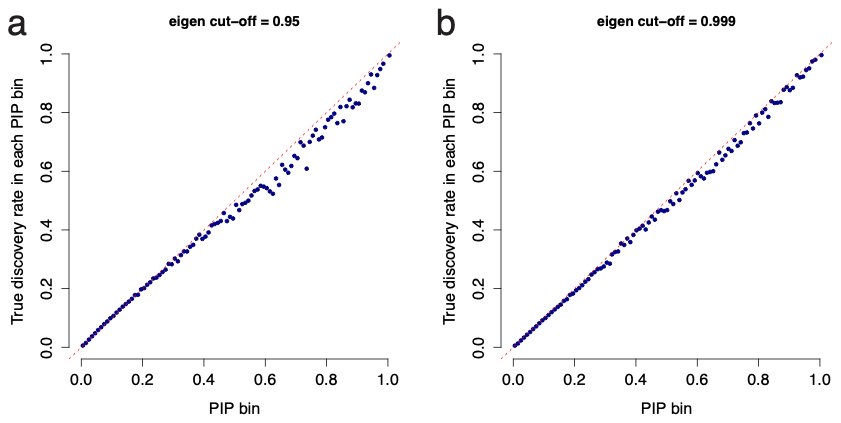

**Supplementary Figure 8** calibration of SBayesRC using different eigen cut-off values under sparse genetic architecture.

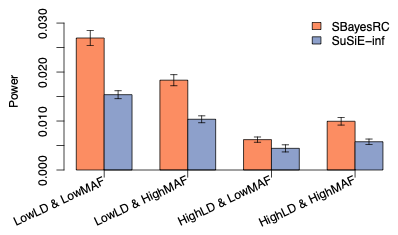

**Supplementary Figure 9** Comparison of power at PIP cut-off of 0.9 between SBayesRC and SuSiE-inf at different LD and MAF bins under sparse genetic architecture simulation. To investigate how GWFM’s improvement varies with MAF and other factors, we stratified the fine-mapping power of both SBayesRC and SuSiE-inf across four bins based on LD score and MAF in our simulation study. Note that in the simulation, following the model of Yang et al, we assumed that the causal effect size is inversely proportional to the heterozygosity of the causal variant: $\beta_{j} \sim N(0, {[2p\left( 1-p \right)]}^{-1}\sigma_{\beta}^{2})$, where p is the MAF. This model assumes an equal amount of genetic variance explained by each causal variant regardless of its MAF, consistent with a model of negative selection17. The results showed that the fine-mapping power was generally higher for causal variants with low LD scores and/or low MAF, given the equal amount of genetic variance explained, as it is easier to identify causal variants with few SNPs in LD (Figure R11). Notably, SBayesRC consistently outperformed SuSiE-inf across all groups, particularly in the high LD and high MAF group. In this scenario, causal effects tend to be smeared on multiple SNPs in LD with the causal variants, making it more challenging to estimate individual SNP effects. This result is consistent with the pronounced advantages of GWFM in the simulations with LDMS genetic architecture, where causal variants were sampled from high LD and high MAF regions (Figure 2 and 3). In conclusion, GWFM can improve SNP effect estimation by leveraging information across SNPs, especially for those with small effect sizes, leading to higher power in identifying causal variants in high LD and high MAF regions.

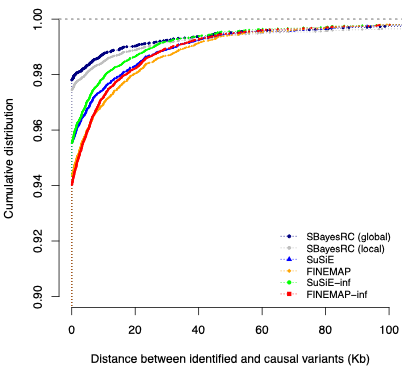

**Supplementary Figure 10** Comparison of different Bayesian methods on distance between the causal variants and identified SNPs using the same either genome-wide or block-wise analysis of SBayesRC and other fine-mapping methods.

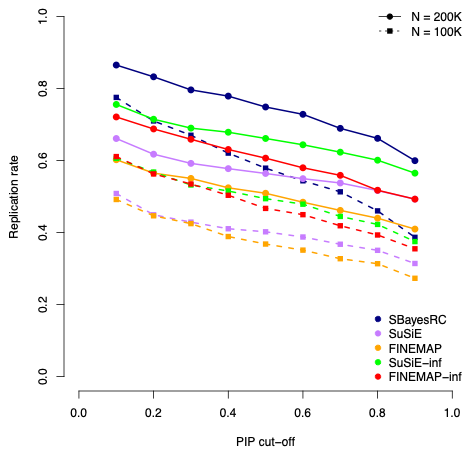

**Supplementary Figure 11** Replication rate of discovery using different methods at a given PIP threshold in the replication sample (x-axis) with different sample sizes for height in the UKB.

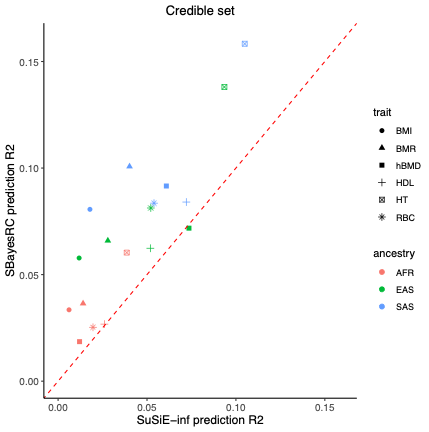

**Supplementary Figure 12** Comparison of trans-ancestry prediction accuracy using fine-mapped variants from SBayesRC and SuSiE-inf from the analysis of samples of European ancestry for 6 complex traits in the UK Biobank, with variants identified in credible sets.

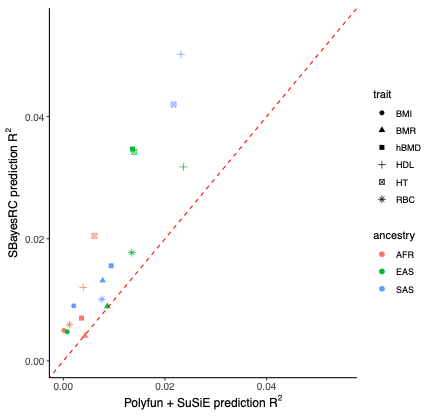

**Supplementary Figure 13** Comparison of trans-ancestry prediction accuracy using fine-mapped variants from SBayesRC and Polyfun + SuSiE from the analysis of samples of European ancestry for 6 complex traits in the UK Biobank.

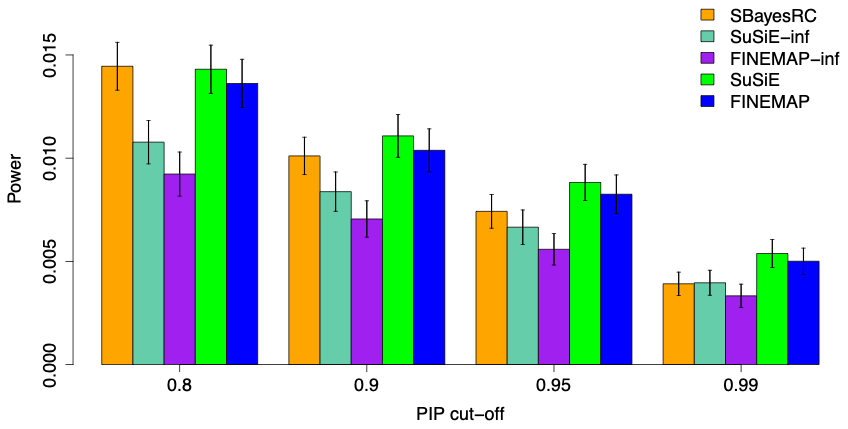

**Supplementary Figure 14** Comparison of power from different methods in the simulation based on the sparse genetic architecture.

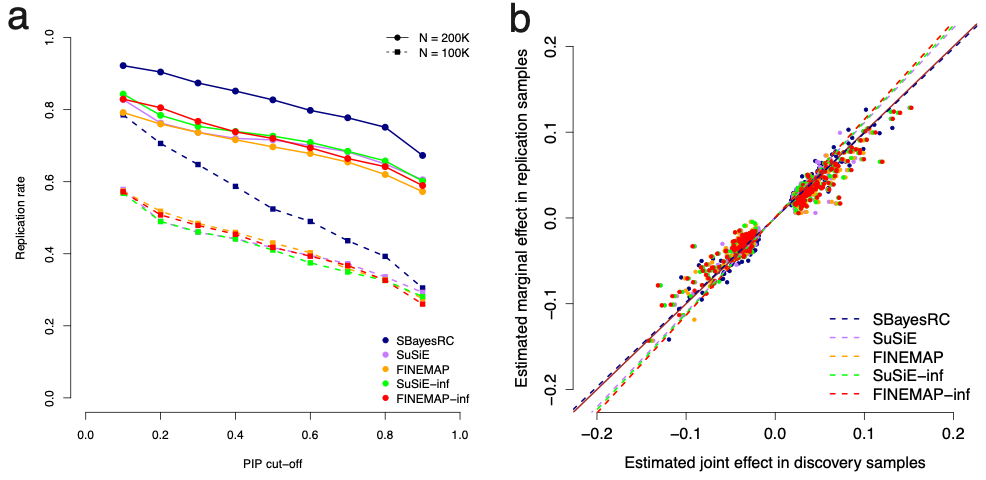

**Supplementary Figure 15** Comparisons of independent sample replication rate, and effect size estimation bias among fine-mapping methods using simulations. Panel a) shows the replication rate of discovery using different methods at a given PIP threshold in the replication sample (x-axis) using simulations. Panel b) shows the regression of the estimated marginal effect size in replication samples on the estimated joint effect size in discovery samples using different fine-mapping methods. Dash line shows the regression slope, which is closer to one for a less biased method. The marginal effect estimated in the independent replication samples was used as a proxy to the true value because it is an unbiased estimate. The brown solid line is the y=x line.

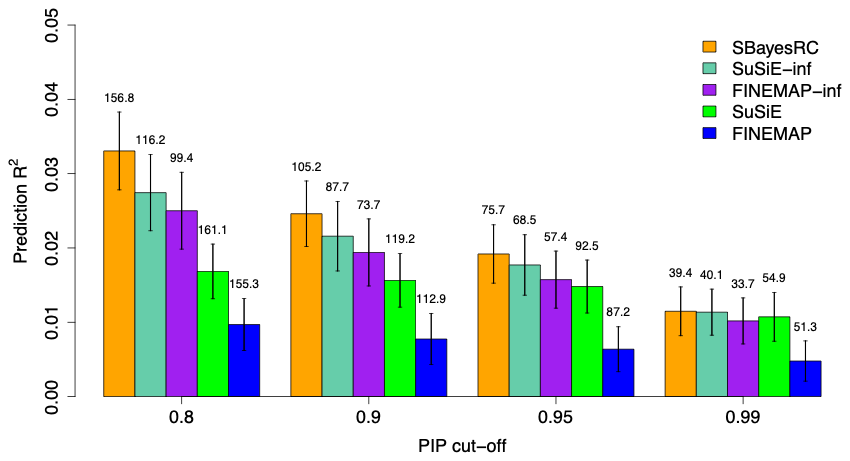

**Supplementary Figure 16** Out-of-sample prediction accuracy using identified variants from different fine-mapping methods. The comparison results for different methods in the simulation is based on the sparse genetic architecture (**Methods**). The number above each bar is the average number of fine-mapped SNPs across 100 simulation replicates from each method at different PIP cut-offs.

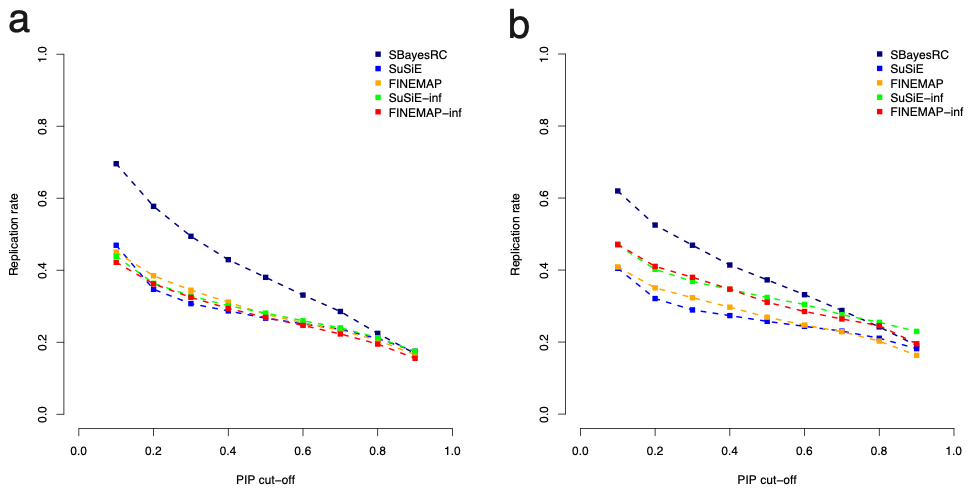

**Supplementary Figure 17** Replication rate of discovery samples of 200,000 using different Bayesian fine-mapping methods at a given PIP threshold in the replication samples of 100,000 (x-axis) using simulations under sparse genetic architecture (a; **Methods**), and using real data analysis for height in the UKB (b).

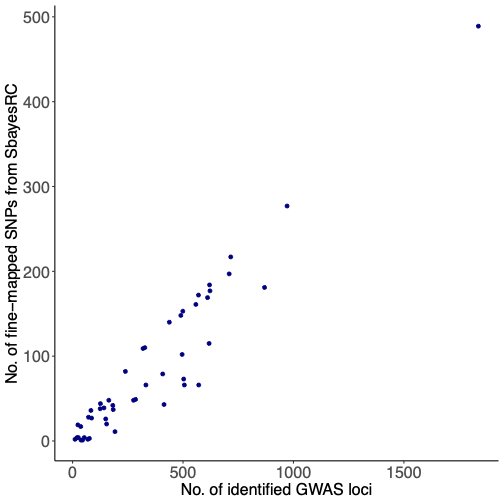

**Supplementary Figure 18** The relationship between the number of fine-mapped variants and the number of identified GWAS loci using LD clumping for 48 complex traits.

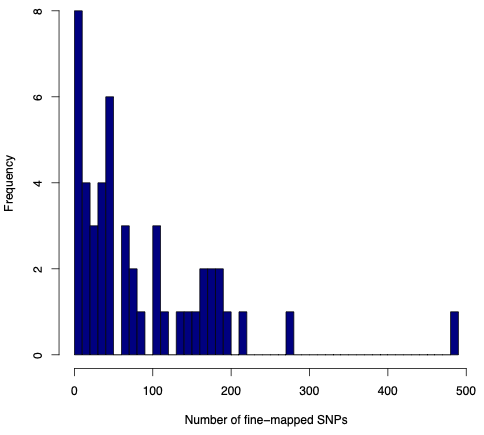

**Supplementary Figure 19** Distribution of the number of fine-mapped SNPs using SBayesRC for 48 complex traits.

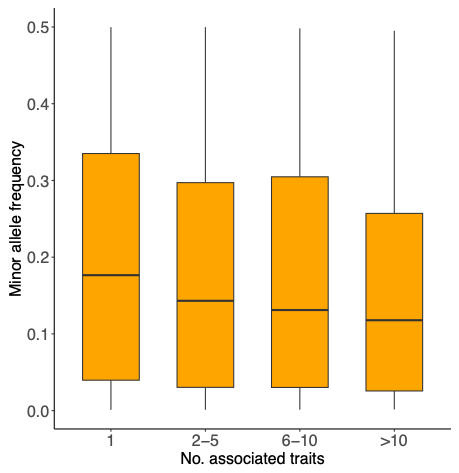

**Supplementary Figure 20** The relationship between the level of pleiotropy (x-axis) and minor allele frequency for fine-mapped SNPs.

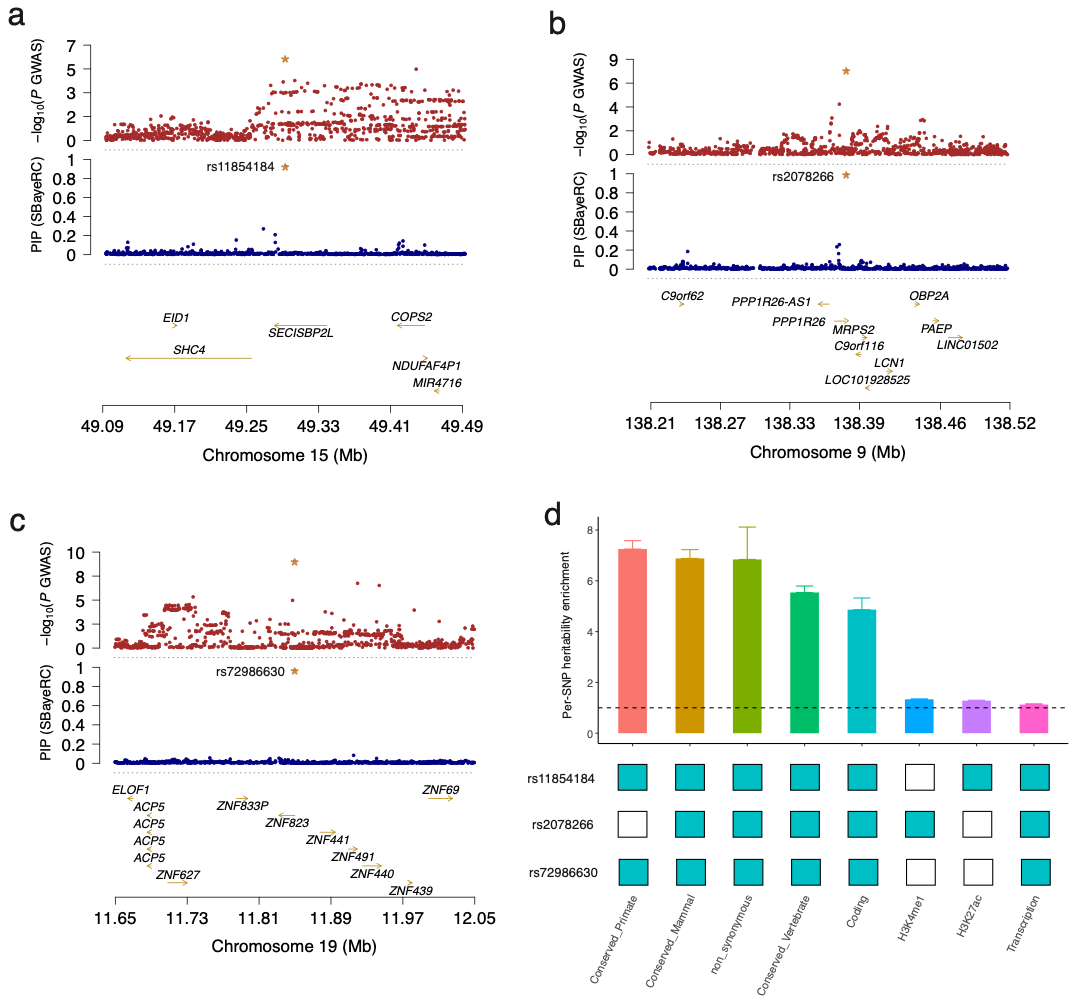
 **Supplementary Figure 21** Prioritized missense variants using SBayesRC fine-mapping analysis for schizophrenia. Panels (a-c) shows the locus plot at the *SECISBP2L*, *PPP1R26* and *ZNF823* locus for schizophrenia, respectively. The top track shows the locus plot of the standard GWAS for schizophrenia, and the bottom track shows the similar plot but with the PIP from SBayesRC for schizophrenia. The starred SNP is the fine-mapped missense variant (i.e., rs11854184, rs2078266 and rs72986630) at each locus, respectively. Panel d shows the heritability enrichment for 8 functional annotations with three fine-mapped missense variants for schizophrenia. The annotations on the x-axis were those present at least once in these three variants, excluding quantitative annotations and duplicated annotations with flanking windows.

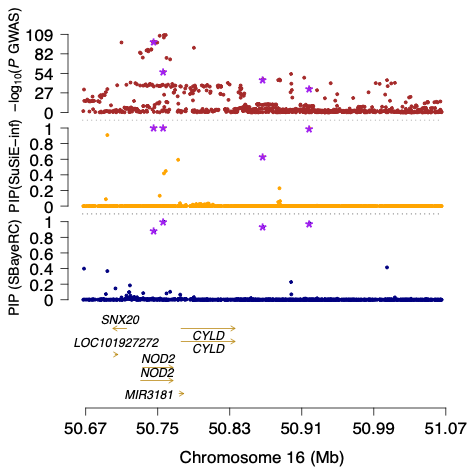

**Supplementary Figure 22** Prioritized causal variant at the *NOD2* locus for Crohn’s disease. The top track shows the *NOD2* locus plot of the standard GWAS for Crohn’s disease, and the second track shows the similar plot but with the PIP from SuSiE-inf, and the third track shows the similar plot but with the PIP from SBayesRC. The starred SNPs are the fine-mapped variants by SBayesRC (rs2066844, rs2066845, rs72798422, and rs145126485).

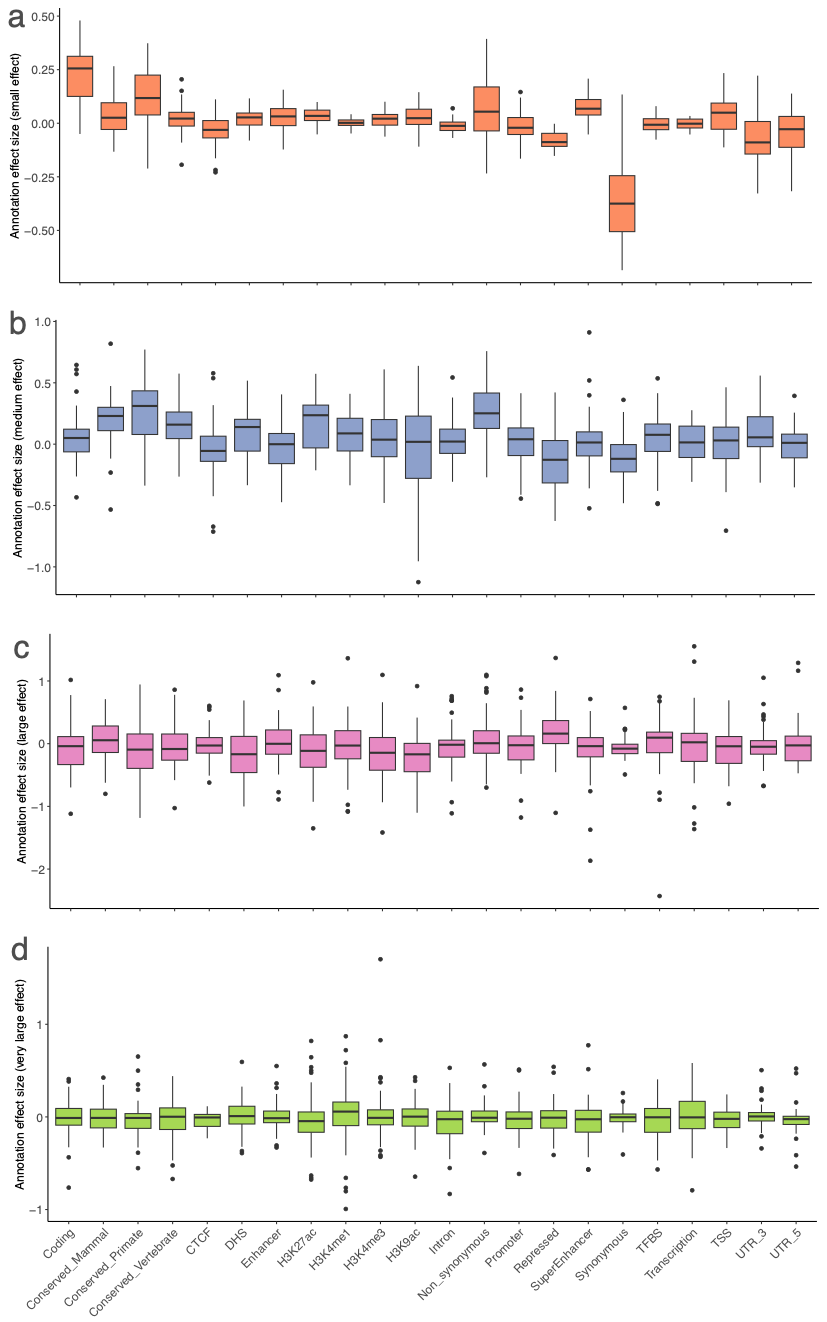

**Supplementary Figure 23** Distribution of annotation effect size for 22 main functional categories for each variance component across 48 complex traits. Panels (a-d) correspond to the “very small”, “small”, “medium”, and “large” effect categories correspond to the four non-zero variance components in SBayesRC, which explain 0.001, 0.01, 0.1, and 1% $h_{SNP}^{2}$, respectively.

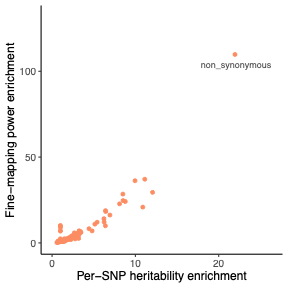

**Supplementary Figure 24** Correlation between fine-mapping power enrichment and per-SNP heritability enrichment in the 97 functional category defined in LDSC baseline model.

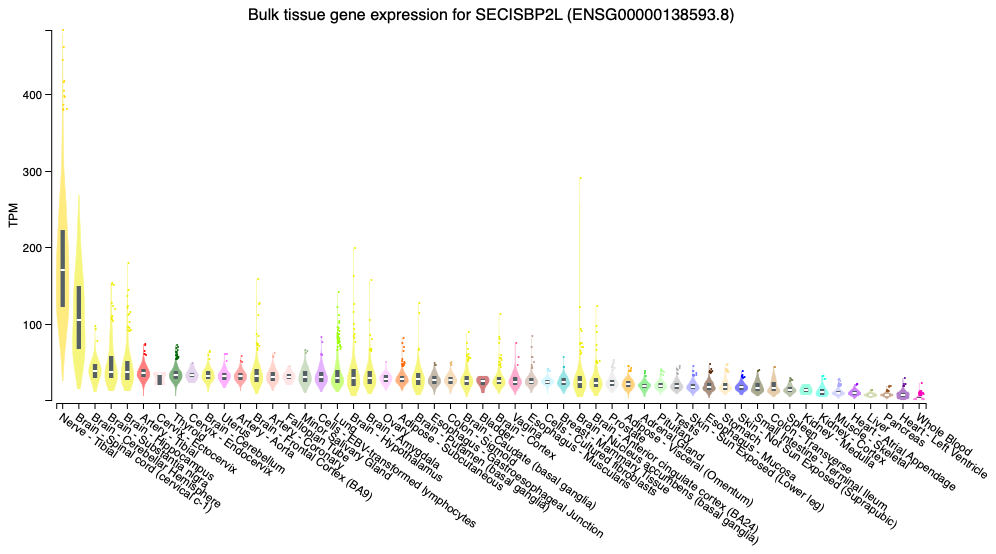

**Supplementary Figure 25** Bulk tissue gene expression for *SECISBP2L* across samples in 49 GTEx tissues.

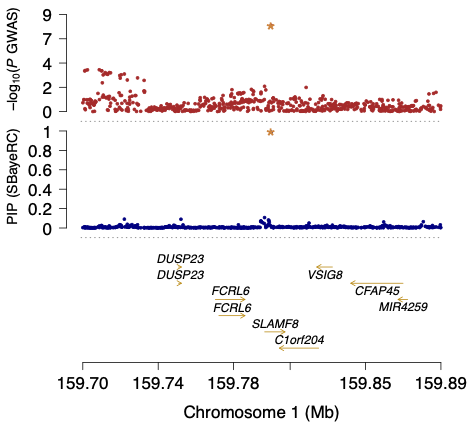

**Supplementary Figure 26** Prioritized missense variants using SBayesRC fine-mapping analysis for Crohn’s disease. The top track shows the locus plot of the standard GWAS for Crohn’s disease, and the bottom track shows the similar plot but with the PIP from SBayesRC for Crohn’s disease. The starred SNP is the fine-mapped missense variant (rs34687326).

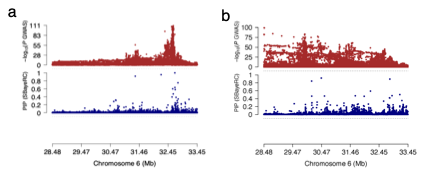

**Supplementary Figure 27** Prioritized causal variant at the MHC locus for asthma (a) and for red blood cell distribution width (b). The top track shows the MHC locus plot of the standard GWAS for asthma and red blood cell distribution width, and the second track shows the similar plot but with the PIP from SBayesRC for asthma and red blood cell distribution width, respectively.

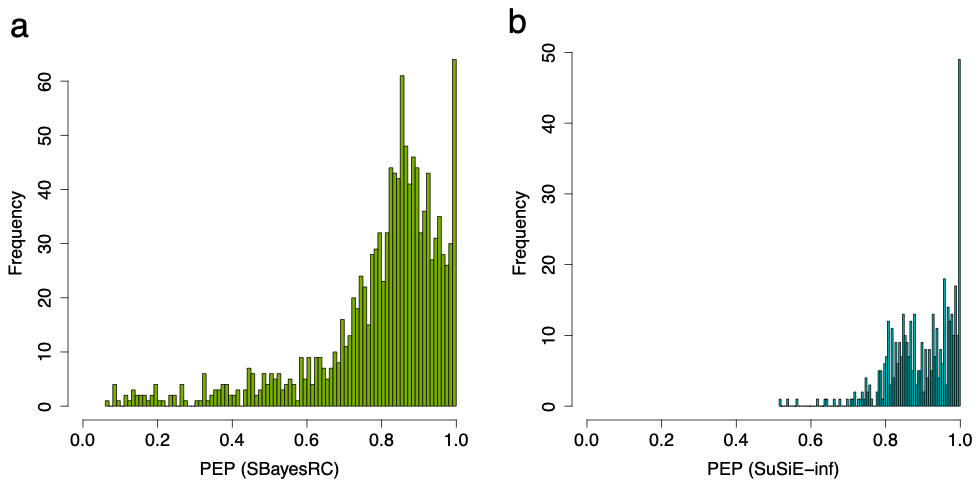

**Supplementary Figure 28** Distribution of posterior enrichment probability (PEP) in SNP-based heritability for local credible sets identified from SBayesRC (a) and SuSiE-inf (b) in simulations with sparse genetic architecture. Since SuSiE-inf reports only posterior mean estimate of SNP effects, PEP cannot be directly computed from its output. Instead, we used the CS SNPs reported by SuSiE-inf while leveraging MCMC samples of their effect sizes from SBayesRC to estimate PEP.

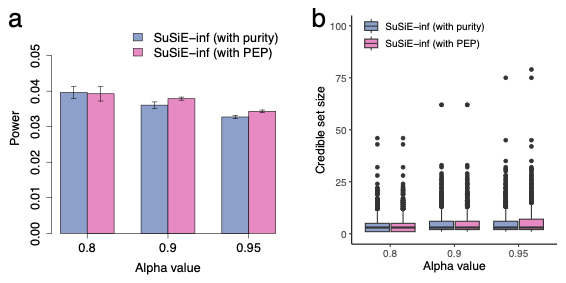

**Supplementary Figure 29** Comparison of power (a) and credible set size (b) for SuSiE-inf with purity filtering or with PEP filtering in simulations with sparse genetic architecture.

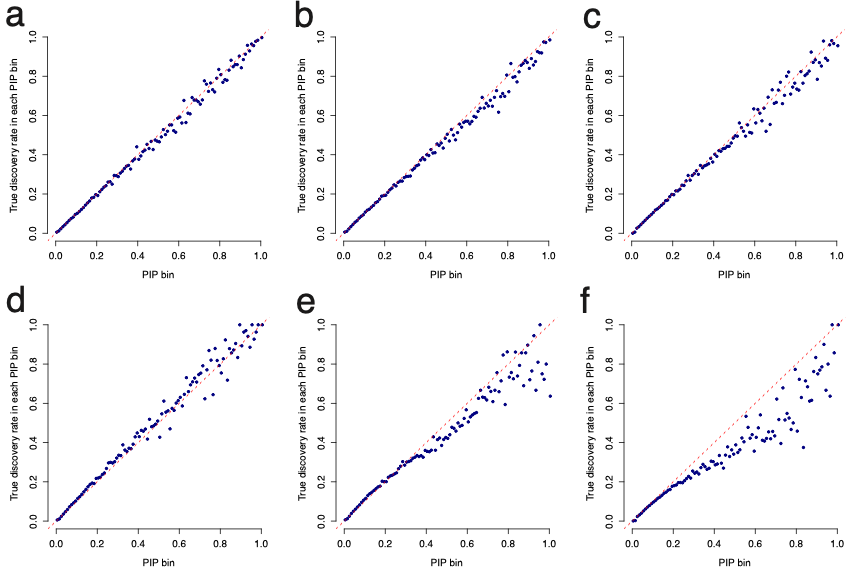

**Supplementary Figure 30** calibration of PIP using an external LD reference of UK10K (a-c) and 1KGP (d-e). Shown are relationship between PIP and the true discovery rate across 100 PIP bins. Results showed in each column correspond to the simulation under sparse genetic architecture (a, and d), large-effects genetic architecture (b and e) and LDMS architecture (c and f).

**Supplementary Table 1** Summary of Bayesian fine-mapping methods used in this study

| **Method** | **Scope** | **Model** | **Algorithm** | **Max no. causals** | **Reference** |
| --- | --- | --- | --- | --- | --- |
| SuSiE | Local | Point-normal mixture | Variational Bayes (VB) | 10 | Wang et al., 2020, J. R. Stat. |
| FINEMAP | Local | Point-normal mixture | Shotgun stochastic search (SSS) | 10 | Benner et al., 2016, Bioinform. |
| SuSiE-inf | Local | Point-normal mixture + polygenic effect | VB | 5 | Cui et al., 2023, Nat. Genet. |
| FINEMAP-inf | Local | Point-normal mixture + polygenic effect | SSS | 5 | Cui et al., 2023, Nat. Genet. |
| SBayesC | Genome-wide | Point-normal mixture | MCMC | Unlimited | Lloyd-Jones et al., 2019, Nat. Comm. |
| SBayesRC | Genome-wide | Multi-normal mixture | MCMC | Unlimited | Zheng et al., 2024, Nat. Genet. |

**Supplementary Table 2** Summary of acronym used in the manuscript

| **Acronym** | **Full name** |
| --- | --- |
| BFP | Body fat percentage |
| BMI | Body mass index |
| BMM | Bayesian mixture model |
| CD | Crohn’s disease |
| COJO | Conditional and joint analysis |
| CS | Credible sets |
| FDR | False discovery rate |
| GBMM | Genome-wide Bayesian mixture model |
| GCS | Global credible sets |
| GWAS | Genome-wide association studies |
| GWFM | Genome-wide fine-mapping |
| HC | Hip circumference |
| HDL | High density lipoprotein |
| ImpLegR | Impedance of leg |
| LCS | Local credible sets |
| LD | Linkage disequilibrium |
| LDMS | LD-and-MAF-stratified |
| MAF | Minor allele frequency |
| MCMC | Markov chain Monte Carlo |
| PEP | Posterior $h_{SNP}^{2}$ enrichment probability |
| PGS | Polygenic scores |
| PHE | Proportion of $h_{SNP}^{2}$ explained by these variants |
| PIP | Posterior inclusion probability |
| SCZ | schizophrenia |
| TDR | True discovery rate |
| TPR | True positive rate |
| UKB | UK Biobank |
| WC | Waist circumference |
