## Supplementary Notes for "Genome-wide fine-mapping improves identification of causal variants"

#### Contents

**1 Comparison of GWFM to existing fine-mapping methods 2**

**2 Sensitivity analyses to assess the thresholds used to construct LCS 4**

**3 Sensitivity analyses to assess the robustness of SBayesRC 4**

**4 Assessing replication rate, effect size estimation, and out-of-sample prediction using fine-mapped variants via simulations 5**

**5 Calculation of SNP PIP 6**

**6 Tempered Gibbs sampling algorithm for sampling SNP indicator variables 7**

**7 Derivation of estimated true positive rate using PIP 8**

**8 Method to predict fine-mapping power 9**

### 1 Comparison of GWFM to existing fine-mapping methods

In contrast to GWAS, which test marginal effects, fine-mapping aims to detect joint-association signals for causal inference, where the strength of joint association is assessed using the posterior inclusion probability (PIP). PIP is the probability of a SNP being included with a nonzero effect in the model, which, in theory, controls false discovery rate (FDR) [1]. Existing methods use similar spike-and-slab mixture model but differ in the algorithm used to derive PIP. For example, FINEMAP [2] utilizes a shotgun stochastic search algorithm to explore possible causal configurations, and computes the PIP by averaging over those with non-negligible probabilities. SuSiE [3] and SuSiE-RSS [4] assume a sparse effect model and employ an iterative Bayesian stepwise selection approach to estimate the overall effect of each SNP by summing up multiple singe-effect vectors. SuSiE-Inf [5] and FINEMAP-Inf [5] further extend the two models to include an infinitesimal component for improved modelling of polygenic architecture within a locus.

The typical fine-mapping strategy focuses on the independently significant GWAS loci (e.g., 1Mb windows centred on lead SNPs with P-value $< 5 \times 10^{-8}$) and performs fine-mapping within these loci separately, assuming at most k (typically $< 10$) causal variants per locus. Our genome-wide fine-mapping (GWFM) approach, SBayesRC, leverages approximately independent LD blocks in the genome and enables "information borrowing" across all blocks to estimate the SNP effect distributions. This strategy improves sensitivity to long-range LD compared to existing methods and accounts for signals over the entire genome. Different to the standard fine-mapping strategy above, our method allows an arbitrary number of causal variants per block, and constructs LCS using posterior samples from the Markov chain Monte Carlo (MCMC) sampling, which is known to be superior to variational inference in accuracy [6, 7].

A high PIP value provides evidence of a causal variant. However, a causal variant may not have a high PIP value if it is in strong LD with other SNPs. For example, if the causal variant is in perfect LD with another SNP, then the PIP is expected to be 0.5 for each variant, given a sufficient sample size. Therefore, the CS concept has been introduced to capture causal variants in strong LD with non-causal SNPs [8, 9]. Our LCS follows the same definition as that in SuSiE – a level of $\alpha$ (local) credible set is a set of SNPs that collectively contain a causal variant with probability $\alpha$. Differences between methods mainly lie in:

1. Selection of SNPs for CS. SuSiE uses a single-effect prior that allows only "exactly one of the p explanatory variables has a non-zero regression coefficient" to inform the credible set, assuming at most $L$ ($\leq$ 10) causal variants. SBayesRC is a genome-wide Bayesian variable selection model which allows arbitrary number of causal variants, estimated from the data. However, while SBayesRC estimates SNP PIP, it does not inherently define how SNPs should be grouped into a credible set. To address this, we propose an LD-based grouping strategy, recognizing that the uncertainty in identifying causal variant arises from LD structure rather than physical distance. Specifically, for a focal SNP with high PIP (candidate causal variant), we identify all SNPs in high LD with the candidate ($r^2 > 0.5$, validated through

simulations) and rank these SNPs in descending order of PIP. We then construct LCS by including SNPs until their cumulative PIP sum reaches or exceeds $\alpha$.

2. Algorithm to estimate SNP PIP. SuSiE employs variational Bayes to approximate the posterior distribution and estimate SNP PIP. In contrast, SBayesRC utilises MCMC sampling, which provides a more accurate posterior distribution by better accounting for uncertainty but lacks methods to define credible sets from MCMC samples. Our work contributes to address this by providing an approach for constructing LCS from MCMC-derived PIPs.

3. CS filtering. The cumulative PIP in a CS will always increase as more SNPs are included. This can lead to overly large CS with many weakly associated SNPs, reducing interpretability and complicating downstream analysis. It is therefore necessary to filter CS to ensure the selected CS are biologically meaningful. SuSiE addresses this issue by applying a purity parameter to filter CS with pairwise absolute LD correlation $< 0.5$, along with a cap of 100 SNPs in a CS. We propose to use posterior enrichment probability (PEP) in SNP-based heritability to ensure the identified CS explains a higher proportion of SNP-based heritability than a random set of SNPs with the same size. This approach enhances interpretability in fine-mapping results.

PEP is conceptually related to the window posterior probability association (WPPA) used in the literature [10], which estimates the posterior probability that the genetic variance explained by a given window exceeds a specified threshold based on MCMC samples. In our study, we introduce PEP to help control the credible set size to ensure the included SNPs are meaningful for fine-mapping. When constructing a credible set, the sum of PIPs will always increase as more SNPs are included. However, this does not necessarily mean that additional SNPs contribute meaningfully to fine-mapping - many could have very small PIPs, making it difficult to distinguish genuine small-effect signals from noise. In contrast, PEP ensures that the SNPs in an LCS collectively explain more genetic variance than a random set of SNPs of the same size. If additional SNPs do not improve this enrichment, PEP will decrease, preventing excessive inclusion of weak associations.

While the concept of credible set has evolved over time [2, 3, 11], it is still common to focus on individual SNP PIP in the downstream analysis, probably because the CS include too many SNPs to follow up. Our study provided important implications regarding this issue. First, CS may miss the true causal variant if not all possible causal variants are fitted in the model, underscoring the importance of considering all common SNPs in the fine-mapping analysis. Second, our GWFM approach can reduce the credible size, as shown in both simulation and real trait analysis (only 5 SNPs per credible set on median), facilitating the consideration of CS in practice. We also investigated whether applying PEP to other fine-mapping methods (i.e., SuSiE-inf) can help improve the performance. Compared to SuSiE-inf with purity filtering, SuSiE-inf with PEP filtering led to slight increase in both the power and credible set size (Supplementary Fig. 29). However, this slight difference may reflect the way

PEP is computed using our MCMC samples. Overall, there was only small difference between SuSiE-inf with purity filtering and with PEP filtering. Third, in the presence of complete LD between SNPs and the causal variants, the PIP of a causal variant may never be significant regardless of sample size, but leveraging functional genomic annotations may help distinguish causal from non-causal variants. In this sense, genomic annotations play a greater role than the increase of GWAS sample size.

### 2 Sensitivity analyses to assess the thresholds used to construct LCS

To assess the robustness of our local credible set (LCS) on the choice of threshold, we ran sensitivity analysis below. We first evaluated the robustness of LCS performance with respect to the choice of posterior $h^2_{SNP}$ enrichment probability (PEP) threshold. To assess the impact of the PEP threshold, we compared power, false discovery rate (FDR), and credible set size (CSS) at different PEP cutoffs, using simulations under various genetic architectures with an $\alpha$-LCS threshold of $\alpha = 0.9$ (Supplementary Fig. 2). We found that the current PEP of 0.7 effectively controls both FDR and CSS, without substantial loss of power. Compared to a PEP threshold of 0.5, using 0.7 yielded a lower FDR with only a small decrease in power, particularly in complex genetic architectures. More importantly, CSS was significantly reduced, improving the resolution and interpretability of fine-mapping. These findings indicate that a PEP threshold of 0.7 provides a balanced performance between power and accuracy, ensuring a more refined set of candidate SNPs without excessive loss of power (Supplementary Fig. 2). However, the "best" choice of PEP (and LD r2) threshold may depend on whether the experimenter values power over the size of the credible set, or the other way around. Therefore, we have made it as an option in our software that can be specified by the user.

We then evaluated the robustness of LCS performance with respect to the choice of LD $r^2$ threshold. To evaluate the impact of LD $r^2$ threshold on GWFM performance, we conducted simulations under the sparse genetic architecture and constructed LCS based on different $r^2$ thresholds. The results showed that while increasing the $r^2$ threshold consistently reduces the credible set size, setting it too high impairs power. Using a threshold of 0.5 provides an optimal balance in reducing the credible set size while maintaining high power (Supplementary Fig. 3).

### 3 Sensitivity analyses to assess the robustness of SBayesRC

We ran a series of sensitivity analyses o assess the robustness of SBayesRC for GWFM. We first assessed the potential impact of missing annotations, we run additional simulations by setting 30% of the total genetic variance from the coding region and 70% from the rest of genome. We ran our SBayesRC analysis with and without coding annotations and found that SBayesRC increased power for causal variants that had correct annotations but slightly missed variants without annotations. (Supplementary

Fig. 6). This suggests that while functional annotations enhance fine-mapping, our method remains robust even when annotation information is incomplete.

To simulate real-world conditions where fine-mapping datasets often do not include the causal variants, we performed additional simulations under a sparse model with 50% of the causal variants missing in the GWAS dataset. Under this scenario, the mapping precision for all methods decreased substantially, whereas our SBayesRC was still superior to the regional-specific methods (Supplementary Fig. 7).

We then investigated the impact of the number of principal components (PCs) retained in the low-rank GBMM. In SBayesRC, the number of PC is automatically determined through pseudo cross-validation within the algorithm. To further validate robustness, we manually tested alternative thresholds (95% and 99.9%) and observed minimal differences in PIP calibration (Supplementary Fig. 8). This finding confirmed that SBayesRC is generally robust to different choices of eigenvalue thresholds.

To understand why GWFM had higher power, we stratified the fine-mapping power of both SBayesRC and SuSiE-inf across four bins based on LD score and MAF. While both methods identified a large proportion of causal variants in the low LD and low MAF group, SBayesRC demonstrates superior performance particularly in the high LD and high MAF groups, highlighting its robustness in complex genomic regions (Supplementary Fig. 9).

To further justify our choice of SBayesRC as the method for GWFM, we ran SBayesRC within each block separately and quantified the mapping precision. We found that the mapping precision decreased compared to the genome-wide SBayesRC but remained higher than the other methods (Supplementary Fig. 10). For example, 99% of SNPs identified by the region-specific SBayesRC were located within 23.1Kb to causal variants, compared to the number of 16.4Kb for the genome-wide SBayesRC and 25.8Kb for SuSiE-inf.

### 4 Assessing replication rate, effect size estimation, and out-of-sample prediction using fine-mapped variants via simulations

We performed simulations using the UKB samples of European ancestry and split samples into independent datasets for discovery and replication. Putative causal variants were identified at the PIP threshold of 0.9 in the GWAS data (n=100,000). We then quantified the replication rate of the putative causal variants at different significance thresholds in two replication datasets (n=100,000 and 200,000). Using SBayesRC, roughly 33% of identified SNPs can be replicated at $PIP > 0.9$ when replication $n = 100,000$, and the replication rate increased to 71% when the replication sample size was doubled (Supplementary Fig. 15a). It may seem counter-intuitive that only a fraction of SNPs was replicated despite using the same PIP threshold of 0.9 in both the discovery and replication datasets. This discrepancy is because there exists a sampling variation in the causal variants identified from distinct samples. As expected, the replication rate increased when using a lower threshold for replication, e.g., with $PIP > 0.1$, 78.8% of the identified SNPs can be replicated when replication $n = 100,000$. Compared to other methods, SBayesRC demonstrated significantly higher replication rate at each of the PIP thresholds, while differences among the other four methods were small.

We then assessed bias in effect size estimates of putative causal variants through regressing their marginal effect sizes from the replication sample on the joint effect sizes estimated from the GWAS sample, expecting a regression slope of one for unbiased estimation. In the simulation results, SBayesRC produced the minimal bias, with a regression slope of 0.97, outperforming all the other methods (Supplementary Fig. 15b). This likely reflects the capacity of SBayesRC to estimate genetic architecture genome-wide, whereas other methods estimate genetic architecture locally or use preset parameters.

To further evaluate the results of fine-mapping, we conducted an out-of-sample prediction using the fine-mapped variants. Since all the Bayesian methods used in this study provide the posterior mean of SNP effects, we computed polygenic scores (PGS) based on the identified variants from each of the methods and evaluated the prediction accuracy in a validation sample. We split the 100K samples into 95K training samples to perform the fine-mapping analysis using all these Bayesian methods and predicted the phenotype for 5K independent individuals as validation samples. We found that overall, SBayesRC had a higher prediction accuracy compared to the other methods, outperforming them by at least $\tilde{1}7\%$ at a PIP threshold of 0.9, with a relatively smaller number of SNPs included in the predictor (Supplementary Fig. 16). This is consistent with the result that SBayesRC resulted in a lower FDR than the other methods (Fig. 2a).

The improved prediction accuracy can be attributed to both higher power and more accurate effect size estimation. As shown in Supplementary Fig. 16, GWFM identified more causal variants than SuSiE-inf, consistent with the observed increased power compared to SuSiE-inf (Supplementary Fig. 16). Moreover, the effect size comparison in Fig. 4b demonstrated that GWFM provided more accurate effect size estimates compared to the other fine-mapping methods (e.g., regression slope of 0.981 from SBayesRC vs. 1.082 from SuSiE-inf), indicating excessive shrinkage in the estimates from the other fine-mapping methods. These findings highlight the key advantages of SBayesRC—its ability to identify more causal variants and provide more precise effect size estimates, both of which contribute to its superior prediction accuracy.

### 5 Calculation of SNP PIP

We assessed the strength of joint association of each SNP using PIP, i.e., the probability of a SNP being included with a nonzero effect in the model, given the data ($\mathbf{w}$). Let $\delta_j$ be the indicator variable for the distribution membership for SNP $j$, with $\delta_j = 1$ indicating a null effect and $\delta_j = 2, \ldots, 5$ indicating a nonzero component. In SBayesRC, we computed PIP for SNP $j$ as

$$PIP_j = \sum_{k=2}^{5} \Pr(\delta_j = k|\mathbf{w}) \quad (1)$$

$$= 1 - \Pr(\delta_j = 1|\mathbf{w}) \quad (2)$$

In the literature, $\Pr(\delta_j = 1|\mathbf{w})$, the probability that SNP $j$ has zero effect is often calculated by counting the frequency of $\delta_j = 1$ in MCMC samples. To improve precision, we use Rao-Blackwellized estimates [12, 13] and compute the posterior mean of $[1 - \Pr(\delta_j = 1|\mathbf{w}, \boldsymbol{\theta})]$ conditional on data and all the other parameters except $\beta_j$ ($\boldsymbol{\theta}$) across $T$ iterations. Suppose $p_j^{(t)} = 1 - \Pr(\delta_j = 1|\mathbf{w}, \boldsymbol{\theta}^{(t)})$, we have

$$PIP_j = E[p_j^{(t)}] \tag{3}$$

$$= \frac{1}{T}\sum_{t=1}^{T}[1 - \Pr(\delta_j = 1|\mathbf{w}, \boldsymbol{\theta}^{(t)})] \tag{4}$$

where

$$\Pr(\delta_j = 1|\mathbf{w}, \boldsymbol{\theta}^{(t)}) = \frac{f(\mathbf{w}|\delta_j = 1, \boldsymbol{\theta}^{(t)})\pi_{j1}}{\sum_{k=1}^{5} f(\mathbf{w}|\delta_j = k, \boldsymbol{\theta}^{(t)})\pi_{jk}} \tag{5}$$

with $f(\mathbf{w}|\delta_j = k, \boldsymbol{\theta}^{(t)})$ being the likelihood function given $\delta_j$ and the sampled values of other parameters except $\beta_j$.

When $\delta_j = 1$, the likelihood function is

$$f(\mathbf{w}|\delta_j = 1, \boldsymbol{\theta}^{(t)}) \propto \exp\{-\frac{\mathbf{w}_c'\mathbf{w}}{2\sigma_e^2}\} \tag{6}$$

where $\mathbf{w}_c = \mathbf{w} - \sum_{j' \neq j} \mathbf{Q}_{j'}\beta_{j'}^{(t)}$ is the adjusted $\mathbf{w}$ for all other effects except that for SNP $j$.

When $\delta_j = k$, the likelihood function is

$$f(\mathbf{w}|\delta_j = k, \boldsymbol{\theta}^{(t)}) \propto \exp -\frac{\mathbf{w}_c'\mathbf{w}}{2\sigma_e^2}\lambda_k^{\frac{1}{2}} C_k^{-\frac{1}{2}} \exp\{\frac{r_j^2}{2C_k}\} \tag{7}$$

where $\lambda_k = \frac{\sigma_e^2}{\gamma_k \sigma_g^2}, C_k = n + \lambda_k$, and $r_j = \mathbf{Q}_{j'}\mathbf{w}_c$.

### 6 Tempered Gibbs sampling algorithm for sampling SNP indicator variables

Joint analysis of all common variants presents a significant challenge for MCMC mixing due to the presence of many variants in strong or complete LD. The standard Gibbs sampling (GS) algorithm can suffer from poor mixing when exploring the joint posterior distribution of variant effects in such cases. For example, in a finite number of iterations, a common causal variant may fail to enter the model if the SNP in complete LD with it has already been selected. To address this issue, we incorporated a tempered Gibbs sampling (TGS) algorithm [14] into our SBayesRC model. Essentially, TGS improves mixing by (1) strategically selecting SNPs for updating ("informed choice") – focusing on those whose indicator variable $\delta_j$ is currently in the tail of the full conditional distribution, and (2) allowing for longer jumps across local maxima by sampling $\delta_j$ from a tempered distribution. Following [14], each MCMC iteration of the algorithm consists of the following key steps:

1. (*SNP selection*) Sample SNP $j$ from $\{1, ..., m\}$ proportionally to $p_{\delta_j} = \frac{1}{K \times \Pr(\delta_j|\mathbf{w}, \boldsymbol{\theta})}$;
2. (*Tempered update*) Sample $\delta_j \sim g_{\delta_j}$;
3. (*Importance weighting*) Weight the new state $\boldsymbol{\delta}$ with a weight $Z(\boldsymbol{\delta})^{-1}$ where $Z(\boldsymbol{\delta}) = \frac{1}{m}\sum_{j=1}^{m} p_{\delta_j}$.

For each SNP, we set the tempered conditional distribution $g_{\delta_j} = g(\delta_j|\delta_{-j})$ to be a Dirichlet distribution with degrees of freedom of one. In step 1, $p_{\delta_j} = p(\delta_j|\mathbf{w}, \boldsymbol{\theta})$ denotes the probability that $\delta_j$ takes its current value conditionally on the current values of all the other parameters and data $\mathbf{w}$. For computational efficiency, we applied TGS only to SNPs with nonzero effects sampled from the standard GS, whose running PIP is greater than 0.01, to assess whether alternative SNPs in high LD ($r^2 > 0.95$) could provide a better fit.

Below shows a pseudocode for the TGS algorithm and a proof-of-concept result from a simple simulation.

**Algorithm 1** Tempered Gibbs sampling for $\delta_j$

**Require:** $\mathbf{w}^{(\mathbf{t})}$ and $\boldsymbol{\theta}^{(t)}$ from $t^{th}$ MCMC iteration

Compute $p_{\delta_j}$ conditional on the sampled values of $\mathbf{w}^{(t)}$ and $\boldsymbol{\theta}^{(t)}$

**for** $i \leftarrow 1$ to $m$ **do**

  Select SNP $j$ based on $p_{\delta_j}$

  Sample new state for $\delta_j \sim Dirichlet(\mathbf{1})$

  Update $\Pr(\delta|\mathbf{w}^{(t,i)}, \boldsymbol{\theta}^{(t,i)})$ for SNPs affected by $\delta_j$

  Compute $p_\delta$ for new state

  Compute weight $Z_i(\boldsymbol{\delta})^{-1}$ for importance sampling

  Sample new effect for SNP $j$: $\beta_j|\delta_j \sim N(\frac{r_j^2}{C_k}, \frac{\sigma_e^2}{C_k})$

**end for**

Update $\Pr(\delta|\mathbf{w}^{(t)}, \boldsymbol{\theta}^{(t)}) = \frac{\sum_{i=1}^{m} \Pr(\delta|\mathbf{w}^{(t,i)}, \boldsymbol{\theta}^{(t,i)}) \times Z_i(\boldsymbol{\delta})^{-1}}{\sum_{i=1}^{m} Z_i(\boldsymbol{\delta})^{-1}}$

### 7 Derivation of estimated true positive rate using PIP

We show below that the genome-wide true positive rate given a posterior in- clusion probability threshold alpha can be estimatd using the PIPs from genome-wide PIPs,

$$\hat{\text{TPR}}(\alpha) = \Pr(\text{PIP} \geq \alpha|\text{H}_1\text{is true}) \tag{8}$$

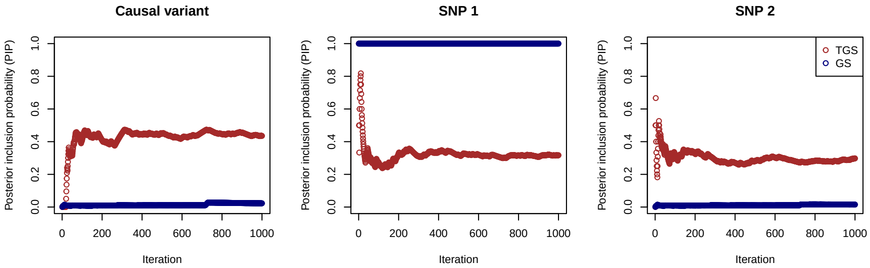

**Fig. 1** Proof-of-concept result demonstrating improved mixing properties of the tempered Gibbs sampler (TGS) compared to the standard Gibbs sampler (GS). Three variants in complete LD were simulated, with only one being causal. GS may assign a posterior inclusion probability (PIP) of 1 to any of the three variants and 0 to the others. In contrast, TGS distributes the PIP more evenly across the three variants, reflecting improved mixing and exploration of the posterior distribution.

$$= \frac{\Pr(\text{PIP} \geq \alpha, \text{H}_1\text{is true})}{\Pr(\text{H}_1\text{is true})} \tag{9}$$

$$= \frac{\Pr(\text{H}_1\text{is true}|\text{PIP} \geq \alpha)\Pr(\text{PIP} \geq \alpha)}{\pi} \tag{10}$$

$$= \frac{\sum_i[\text{PIP}_i|\text{PIP}_i \geq \alpha]}{\#\{\text{PIP}_i \geq \alpha\}} * \frac{\#\{\text{PIP}_i \geq \alpha\}}{\text{M}} * \frac{1}{\pi} \tag{11}$$

$$= \frac{\sum_i[\text{PIP}_i|\text{PIP}_i \geq \alpha]}{\text{M}\pi} \tag{12}$$

where M is the total number of SNPs, and $\pi$ is the proportion of SNPs with nonzero effects on complex trait. One underlying assumption for our derivation above is $\Pr(\text{H}_1\text{is true}|\text{PIP} \geq \alpha) = \text{PIP}$, which has been shown is true in Figure 2 of our main text.

When considering each focal SNP $j$, we can also compute its corresponding true positive rate with the genome-wide PIPs by replacing the threshold $\alpha$ with an observed $\text{PIP}_j$,

$$\hat{\text{TPR}}_j = \frac{\sum_i[\text{PIP}_i|\text{PIP}_i \geq \text{PIP}_j]}{\text{M}\pi} \tag{13}$$

### 8 Method to predict fine-mapping power

We aim to develop a method to predict the power of fine-mapping using a genome-wide Bayesian mixture model (GBMM). The goal is to find the required sample size for identifying 80% of the causal variants given a PIP threshold of 0.9.

For each SNP, it is assumed that

$$\beta_j \sim \sum_{k=1}^{5} \pi_k N(0, \gamma_k \sigma_g^2) \tag{14}$$

Let $\delta_j$ be the indicator variable for the distribution membership for SNP $j$. The posterior inclusion probability (PIP) is

$$\text{PIP} = 1 - \Pr(\delta_j = 1|y) \tag{15}$$

For notation brevity, we will ignore the subscript $j$ in the following derivation, as it is generic to any SNP.

#### 8.1 Distribution of PIP

$$\begin{aligned}\text{PIP} &= 1 - \frac{f(y|\delta=1)\pi_1}{\sum_{k=1}^{5} f(y|\delta=k)\pi_k} &(16)\\ &= 1 - \frac{1}{1+\sum_{k=2}^{5}\frac{f(y|\delta=k)}{f(y|\delta=1)}\frac{\pi_k}{\pi_1}} &(17)\end{aligned}$$

The likelihood functions are

$$f(y|\delta=1) \propto \exp\{-\frac{y_c' y_c}{2\sigma_e^2}\} \tag{18}$$

where $y_c$ is the adjusted $y$ for all other effects except $\beta$.

$$\begin{aligned}f(y|\delta=k) &\propto \int f(\beta|\delta)f(y|\delta=k,\beta)\mathrm{d}\beta &(19)\\ &\propto \exp\{-\frac{y_c' y_c}{2\sigma_e^2}\}(\gamma_k\sigma_g^2)^{-\frac{1}{2}}(C_k^{-1}\sigma_e^2)^{\frac{1}{2}}\exp\{\frac{1}{2}rC_k^{-1}r\} &(20)\\ &\propto \exp\{-\frac{y_c' y_c}{2\sigma_e^2}\}\lambda_k^{\frac{1}{2}}C_k^{-\frac{1}{2}}\exp\{\frac{r^2}{2C_k}\} &(21)\end{aligned}$$

where

$$\begin{aligned}\lambda_k &= \frac{\sigma_e^2}{\gamma_k\sigma_g^2} &(22)\\ C_k &= n+\lambda_k &(23)\\ r &= X'y_c = X'(e+X\beta) = X'e+n\beta &(24)\end{aligned}$$

Therefore,

$$\frac{f(y|\delta=k)}{f(y|\delta=1)} = \lambda_k^{\frac{1}{2}}C_k^{-\frac{1}{2}}\exp\{\frac{r^2}{2C_k}\} \tag{25}$$

and

$$\sum_{k=2}^{5}\frac{f(y|\delta=k)}{f(y|\delta=1)}\frac{\pi_k}{\pi_1} = \sum_{k=2}^{5}\frac{\pi_k}{\pi_1}\lambda_k^{\frac{1}{2}}C_k^{-\frac{1}{2}}\exp\{\frac{r^2}{2C_k}\} \tag{26}$$

In this equation, only $r$ is a random variable that depends on $y$. Given the true $\beta$ value,

$$\mathrm{E}[r] = n\beta \tag{27}$$
$$\mathrm{Var}[r] = n\sigma_e^2 \tag{28}$$

Therefore,

$$r \sim N(n\beta, n\sigma_e^2) \tag{29}$$

Let $v = \beta^2$ be the variance explained by the SNP, then we have

$$r^2 \sim n\sigma_e^2 \chi_1^2(\frac{nv}{\sigma_e^2}) \tag{30}$$

Let

$$Z = \chi_1^2(\frac{nv}{\sigma_e^2}) \tag{31}$$

Substituting into (26) gives

$$\sum_{k=2}^{5} \frac{f(y|\delta = k)}{f(y|\delta = 1)} \frac{\pi_k}{\pi_1} = \sum_{k=2}^{5} \frac{\pi_k}{\pi_1} \lambda_k^{\frac{1}{2}} C_k^{-\frac{1}{2}} \exp\{\frac{n\sigma_e^2}{2C_k} Z\} \tag{32}$$

Let

$$A_k = \frac{\pi_k}{\pi_1} \lambda_k^{\frac{1}{2}} C_k^{-\frac{1}{2}} \tag{33}$$
$$B_k = \frac{n\sigma_e^2}{2C_k} \tag{34}$$

Then,

$$\mathrm{PIP} = 1 - \frac{1}{1 + \sum_{k=2}^{5} A_k \exp\{B_k Z\}} \tag{35}$$

It shows that PIP is a function of a non-central Chi-square variable with NCP $= \frac{nv}{\sigma_e^2}$.

#### 8.2 Analytic solution for two-component mixture

If the number of mixture components $K = 2$, then a point-normal mixture is assumed,

$$\beta \sim \pi N(0, \sigma_\beta^2) + (1 - \pi)\phi_0 \tag{36}$$

In this case,

$$\mathrm{PIP} = 1 - \frac{1}{1 + A \exp\{BZ\}} \tag{37}$$

where

$$A = \frac{\pi}{1 - \pi} \lambda^{\frac{1}{2}} C^{-\frac{1}{2}} \tag{38}$$

$$B = \frac{n\sigma_e^2}{2C} \tag{39}$$

For notation brevity, let P = PIP. Rearranging (37) gives

$$Z = \frac{1}{B}(\log P - \log(1-P) - \log A) \tag{40}$$

$$= u(P) \tag{41}$$

The derivative of $u(P)$ with respect to P is

$$\frac{\partial u(P)}{\partial P} = \frac{1}{B}(\frac{1}{P} + \frac{1}{1-P}) = \frac{1}{B}\frac{1}{P(1-P)} \tag{42}$$

Therefore, the probability density function of PIP is

$$f(P) = f_Z(u(P))|\frac{\partial u(P)}{\partial P}| \tag{43}$$

$$= \text{dchisq}(u(P), 1, \frac{nv}{\sigma_e^2})\frac{1}{B}\frac{1}{P(1-P)} \tag{44}$$

#### 8.3 Analytic solution for multi-component mixture

When $K > 2$, e.g., $K = 5$, rearranging (35) gives

$$\sum_{k=2}^{5} A_k \exp\{\sum_{k=2}^{5} B_k Z\} = \frac{P}{1-P} \tag{45}$$

In recognition of $P = \sum_{k=2}^{5} P_k$, the above equation can break down to be the following set of equations.

$$A_k \exp\{B_k Z\} = \frac{P_k}{1-P}, \quad \text{for } k = 2, ..., 5 \tag{46}$$

Taking the product of the set of equations gives

$$\prod_{k=2}^{5} A_k \exp\{B_k Z\} = \prod_{k=2}^{5} \frac{P_k}{1-P} \tag{47}$$

Taking the logorithm on both sides gives

$$\sum_{k=2}^{5} \log A_k + \sum_{k=2}^{5} B_k Z = \sum_{k=2}^{5} \log P_k - 4\log(1-P) \tag{48}$$

Rearranging the equation gives

$$Z = \frac{\sum_{k=2}^{5} \log P_k - 4\log(1-P) - \sum_{k=2}^{5} \log A_k}{\sum_{k=2}^{5} B_k} \tag{49}$$

However, we don't know individual value of $P_k$. As a proxy, we set

$$P_k = P\frac{\pi_k}{\sum_{k=2}^{5} \pi_k} \tag{50}$$

Therefore,

$$Z = \frac{4\log P + \sum_{k=2}^{5} \log \pi_k - 4\log(1-\pi_1) - 4\log(1-P) - \sum_{k=2}^{5} \log A_k}{\sum_{k=2}^{5} B_k} \tag{51}$$

$$= u(P) \tag{52}$$

The derivative of $u(P)$ with respect to $P$ is

$$\frac{\partial u(P)}{\partial P} = \frac{4}{\sum_{k=2}^{5} B_k} \frac{1}{P(1-P)} \tag{53}$$

Then,

$$f(P) = f_Z(u(P))|\frac{\partial u(P)}{\partial P}| \tag{54}$$

$$= \text{dchisq}(u(P), 1, \frac{nv}{\sigma_e^2})\frac{4}{\sum_{k=2}^{5} B_k}\frac{1}{P(1-P)} \tag{55}$$

#### 8.4 Power calculation

Given a GWAS sample size, we are interested in the power for identifying the causal variants. This will, in turn, predict the required sample size to achieve a certain level of power.

Conditional on a per-SNP variance explained $v$, when a positive result is claimed at the PIP threshold of 0.9, the power can be calculated as

$$\text{Power}_v = \Pr(PIP > 0.9|v) \tag{56}$$

$$= \int_{0.9}^{1} f(P|v)\mathrm{d}P \tag{57}$$

To compute the power for identifying any causal variant, we need to further integrate out $v$:

$$\text{Power} = \int_{0.9}^{1}\int_{0}^{\infty} f(P|v)f(v)\mathrm{d}v\mathrm{d}P \tag{58}$$

Since $v = \beta^2$, then $\beta = v^{\frac{1}{2}} = u(v)$. The distribution of $v$ is

$$f(v) = f_\beta(u(v))2|u'(v)| \tag{59}$$
$$= f_\beta(v^{\frac{1}{2}})v^{-\frac{1}{2}} \tag{60}$$

where $f_\beta$ is the distribution of $\beta$ which is a mixture of normal distributions.

To compute the power for identifying causal variants that altogether explain $\rho$% genetic variance, we first find the $(1-\rho)$% quantile of $v$ distribution, $\alpha_v$, and then compute

$$\text{Power} = \int_{0.9}^{1}\int_{\alpha_v}^{\infty} f(P|v)f(v)\mathrm{d}v\mathrm{d}P \tag{61}$$

#### 8.5 Calculation of expected number of fine-mapped causal variants

Given a sample size, the expected number of fine-mapped causal variants (NCV) is

$$\text{NCV} = \sum_{j=1}^{m_c}\int_{0.9}^{1}\int_{\alpha_v}^{\infty} f(P|v)f(v)\mathrm{d}v\mathrm{d}P \tag{62}$$
$$= m(1-\pi_1) \times \text{Power} \tag{63}$$

#### 8.6 Calculation of expected proportion of variance explained

Given a set of identified variants, we are interested in the expected variance explained by these variants

$$\text{E[VE]} = \text{NCV} \times \text{E}[v|P > 0.9] \tag{64}$$
$$= \text{NCV} \times \int_0^\infty vf(v|P > 0.9)\mathrm{d}v \tag{65}$$

where

$$f(v|P > 0.9) = \frac{f(v, P > 0.9)}{f(P > 0.9)} \tag{66}$$
$$= \frac{f(P > 0.9|v)f(v)}{f(P > 0.9)} \tag{67}$$
$$= \frac{f(P > 0.9|v)f(v)}{\int_0^\infty f(P > 0.9|v)f(v)\mathrm{d}v} \tag{68}$$
$$= \frac{\text{Power}_v \times f(v)}{\text{Power}} \tag{69}$$

Therefore,

$$\mathrm{E}[\mathrm{VE}] = m(1-\pi_1)\int_0^{\infty} \mathrm{Power}_v \times v f(v)\mathrm{d}v \tag{70}$$

The expected proportion of variance explained is

$$\mathrm{E}[\mathrm{PVE}] = \frac{m(1-\pi_1)\int_0^{\infty} \mathrm{Power}_v \times v f(v)\mathrm{d}v}{m(1-\pi_1)\int_0^{\infty} v f(v)\mathrm{d}v} \tag{71}$$

$$= \frac{\int_0^{\infty} \mathrm{Power}_v \times v f(v)\mathrm{d}v}{\int_0^{\infty} v f(v)\mathrm{d}v} \tag{72}$$

#### 8.7 Monte Carlo integration

It is challenging to work out the double integral in the above equations. Alternatively, we can estimate the power using Monte Carlo integration. The process is as follows.

First, we draw 10,000 samples of $\beta_j$ based on the estimated mixture distribution of SNP effects (Eq (14)) and compute $v_j = \beta_j^2$. For each $v_j$, we draw 1,000 samples from the non-central chi-square distribution (Eq (31)) and compute $P_k|v_j$.

Then, the power to detect $v_j$ in Eq (58) can be computed as

$$\mathrm{Power}_{v_j} = \frac{1}{1000}\sum_{k=1}^{1000} I(P_{k|v_j} > 0.9) \tag{73}$$

where $I(\cdot)$ is an indicator function gives 1 if true, otherwise gives 0.

The power to detect any causal variant in Eq (61) can be computed as

$$\mathrm{Power} = \frac{1}{10000}\sum_{j=1}^{10000} \mathrm{Power}_{v_j} \tag{74}$$

The expected proportion of variance explained is

$$\mathrm{E}[\mathrm{PVE}] = \frac{\sum_{j=1}^{10000} v_j \times \mathrm{Power}_{v_j}}{\sum_{j=1}^{10000} v_j} \tag{75}$$

#### 8.8 Accounting for LD between causal and non-causal SNPs

The derivation above assumes that the causal variant can be distinguished from the non-causal variants as the sample size increases. However, this would not be the case if the causal variant is in complete LD with some SNPs. To model this, we assume that the number ($l$) of SNPs confounded (e.g., in complete LD) with the causal variant follows a Poisson distribution with mean $\kappa$:

$$l \sim Poisson(\kappa) \tag{76}$$

In this case, the sampling distribution of PIP becomes

$$f(PIP) = \frac{1}{l+1} f(PIP|v) + \frac{l}{l+1} f(PIP|Null) \tag{77}$$

where $f(PIP|Null)$ is the distribution under the null ($v = 0$).

#### 8.9 Numerical integration for power calculation

The double integral in equation (58) can be computed numerically after being rewritten to increase the stability of the numerical integration:

$$\int_{0.9}^{1} \int_{0}^{\infty} f(P|v)\, f(v)\, dv\, dP = \int_{u(0.9)}^{\infty} \int_{0}^{\infty} f(z|v)\, f(v)\, dv\, dz \tag{78}$$

Note that $u(P)$ cannot be expressed in closed form but can be easily computed by dichotomy since $u$ is monotonic.

Let $\lambda$ be the non-centrality parameter: $\lambda = \frac{nv}{\sigma_e^2}$, then

$$\begin{aligned}
f(z|v) &= \text{dchisq}(z, 1, \lambda) && (79)\\
&= \frac{1}{2} e^{-(z+\lambda)/2} \left(\frac{z}{\lambda}\right)^{-1/4} I_{-1/2}(\sqrt{\lambda z}) && (80)\\
&= \frac{1}{2} e^{-(z+\lambda)/2} \left(\frac{z}{\lambda}\right)^{-1/4} e^{\sqrt{\lambda z}} e^{-\sqrt{\lambda z}} I_{-1/2}(\sqrt{\lambda z}) && (81)\\
&= \frac{1}{2} e^{-(z-2\sqrt{\lambda z}+\lambda)/2} e^{-\sqrt{\lambda z}} I_{-1/2}(\sqrt{\lambda z}) && (82)\\
&= \frac{1}{2} e^{-(\sqrt{z}+\sqrt{\lambda})^2/2} e^{-\sqrt{\lambda z}} I_{-1/2}(\sqrt{\lambda z}) && (83)
\end{aligned}$$

where $I_\nu(y)$ is a modified Bessel function of the first kind and we compute $e^{-y} I_\nu(y)$ to avoid overflow when $y$ is large.

Furthermore,

$$f(v) = \sum_{k=2}^{k=K} \frac{\pi_k}{1-\pi_1} \text{dnorm}(\sqrt{v}, 0, \sigma_k^2) v^{-1/2} \tag{84}$$

and (78) becomes

$$\text{Power} = \sum_{k=2}^{k=K} \frac{\pi_k}{1-\pi_1} \int_{u(0.9)}^{\infty} \int_{0}^{\infty} \text{dchisq}(z, 1, \lambda)\, \text{dnorm}(\sqrt{v}, 0, \sigma_k^2)\, v^{-1/2}\, dz\, dv \tag{85}$$

Finally, we proceed to the change of variable $y = \ln(\lambda) = \ln(\frac{nv}{\sigma_e^2})$ to obtain:

$$\text{Power} = \sum_{k=2}^{k=K} \frac{\pi_k}{1-\pi_1} \int_{u(0.9)}^{\infty} \int_{-\infty}^{\infty} \text{dchisq}(z, 1, e^y)\, \text{dnorm}(\sqrt{v}, 0, \sigma_k^2)\, \sqrt{v}\, dz\, dy \tag{86}$$

with $v = \frac{\sigma_e^2}{n} e^y$.
